# Weakening of the cognition and height association from 1957 to 2018: findings from four British birth cohort studies

**DOI:** 10.1101/2021.12.10.21267601

**Authors:** David Bann, Liam Wright, Neil M Davies, Vanessa Moulton

## Abstract

**Background:** On average taller individuals have been repeatedly found to have higher scores on cognitive assessments, yet it is unclear whether the magnitude of this association has systematically changed across time. Recent studies have found that this association can be explained by genetic factors, yet this does not preclude the influence of environmental or social factors that affect the genome. We tested whether the association between cognition and height has weakened across time.

**Methods:** We used four British birth cohorts (born 1946c, 1958c, 1970c, and 2001) with comparable data available at 10/11 and 14/17 years (N = 41,418). Height was measured at each age, and cognition via verbal reasoning (10/11 years) and vocabulary/comprehension scales (14/16 years) and via mathematical tests at both ages. We constructed age-adjusted height and cognition measures and converted cognition measures to ridit scores to aid interpretation. We then used linear and quantile regression to investigate whether cross-sectional associations between cognition and height differed in each cohort, sequentially adjusting for sex, childhood socioeconomic position, and maternal and paternal height.

**Results:** Taller participants had higher mean cognitive assessment scores in childhood and adolescence, yet the associations were weaker in later (1970c and 2001c) cohorts – after adjustment for sex the mean difference in height comparing the highest with lowest verbal cognition scores at 10/11 years was 0.57 SD (95% CI = 0.44, 0.7) in the 1946c, 0.59 SD (0.52, 0.65) in the 1958c, 0.47 SD (0.41, 0.53) in the 1970c, 0.30 SD (0.23, 0.37) in the 2001c. This pattern of change in association was observed across all specifications (ages 10/11 and 14/16 years, and for each cognition measure used), and was robust to adjustment for social class and parental height, and modelling of plausible missing-not-at-random scenarios. Quantile regression suggested that these average differences were driven by differences in the lower centiles of height. This pattern was most evident in older cohorts – for example, in 1958c, the difference in height was 0.73 SD (0.64, 0.82) at the 10th centile, yet 0.46 SD (0.34, 0.57) at the 90th centile.

**Conclusion:** Associations between height and cognitive assessment scores in childhood-adolescence weakened by at least half from 1957 to 2018. These results support the notion that environmental and social change can markedly weaken associations between cognition and other traits.

## Introduction

Cognitive ability is a potentially important determinant of health, with associations between higher cognition and favourable subsequent health outcomes repeatedly documented in childhood, adolescence, and across adulthood.^1^ This body of evidence—termed by some as ‘cognitive epidemiology’—includes associations of lower cognition and anthropometric measures such as shorter height.^2 3^ Shorter height is in turn associated with a range of disease outcomes and, like cognitive development, is influenced by early life factors.^4-7^

Recent studies have suggested that links between higher cognition and taller height can be explained by genetics,^2 3 8 9^ yet this does not preclude the importance of the environment. Indeed, while height is highly heritable (approximately 80%),^10^ substantial increases in average height occurred in the 20^th^ century,^11 12^ likely due to improved nutrition.^6^ Cognitive test scores have also increased in the 20^th^ century (the ‘Flynn’ effect),^13^ suggesting that both factors are responsive to environmental change. Improvements in nutrition and other shared developmental determinants of cognition and height such as infectious disease^5 6^ may have weakened links between cognition and height.^14^ This has been suggested in the analysis of subsample of eastern Danish male conscripts (1939-1967),^15^ yet data from more heterogeneous and contemporary samples is required to strengthen conclusions and aid generalizability.

We conducted a cross-cohort study to investigate if the associations of cognition with child-adolescent height have systematically changed across time (1957-2018). We used four British birth cohort studies, each containing prospectively ascertained assessments of cognition and height. We hypothesized that improvements in the shared determinants of both cognition and height (e.g., the familial environment) would lead to weakening of associations across time; see Supplementary Figure S1 for a causal diagram. We also hypothesized that cognition-height associations would be strongest amongst those in the lowest height centiles, where environmental factors may be particularly influential and the environment most heterogenous.

## Methods

### Study Samples

Four birth cohort studies conducted in Britain with participants followed-up in childhood and adolescence were used. These were born in 1946 (MRC National Survey of Health and Development, 1946c), 1958 (National Child Development Study, 1958c), 1970 (British Cohort Study, 1970c), and 2000/02 (Millennium Cohort Study, 2001c); further details are provided in the cohort profiles.^16-19^ To aid comparability, analyses were restricted to singleton births born in England, Scotland, and Wales. Sample sizes for analyses, in each cohort and age are shown in Table 1. Each study and sample sweep used in this manuscript has received approved ethical approval and obtained parental/participant according to guidance which was in place at the time of data collections from 1957 to 2018. Detailed breakdowns of these processes are provided elsewhere.^20-22^

**Table 1.**
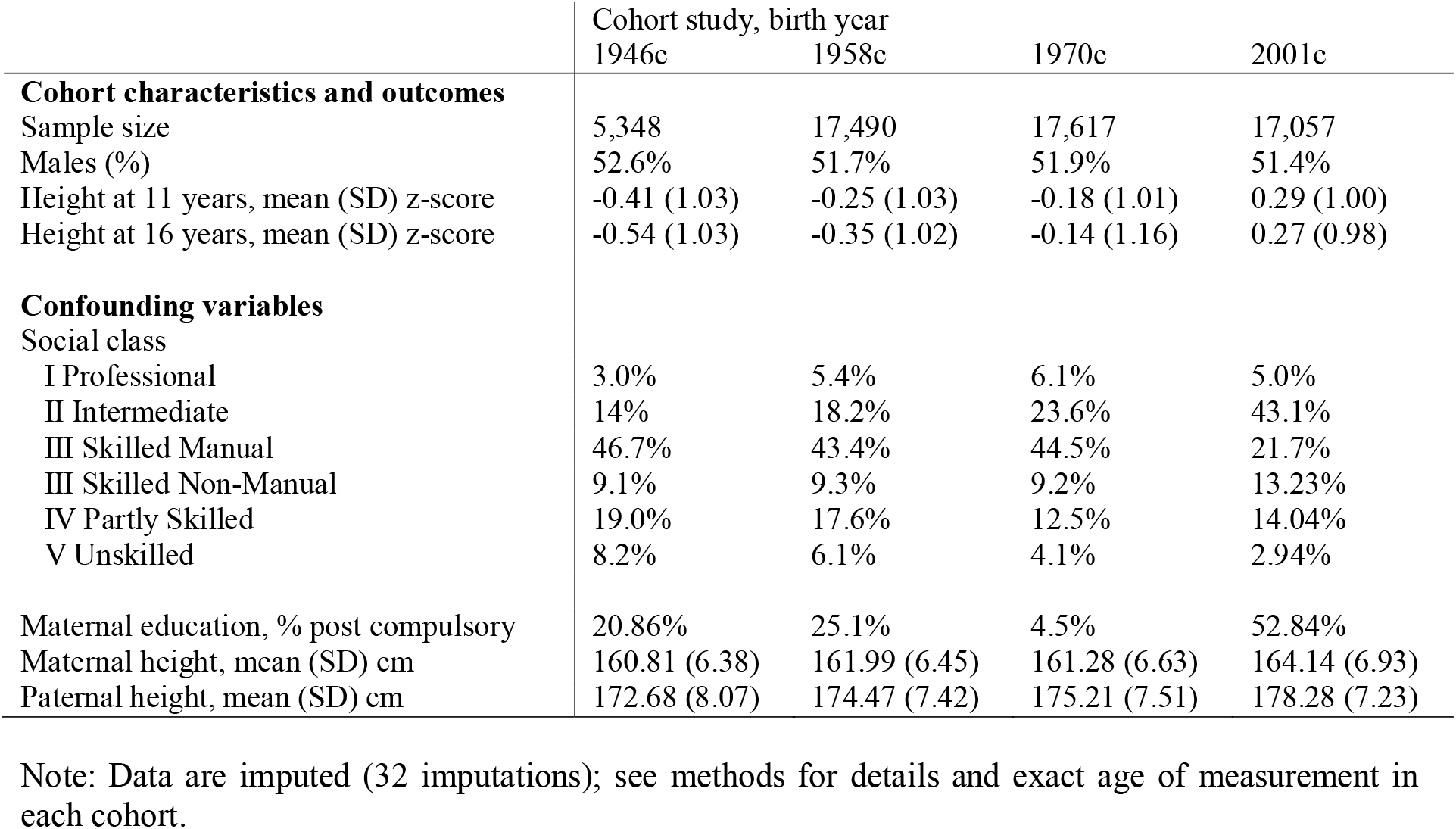
Participant characteristics: data from four British cohort studies.

|  | Cohort study, birth year |  |  |  |
| --- | --- | --- | --- | --- |
|  | 1946c | 1958c | 1970c | 2001c |
| **Cohort characteristics and outcomes** |  |  |  |  |
| Sample size | 5,348 | 17,490 | 17,617 | 17,057 |
| Males (%) | 52.6% | 51.7% | 51.9% | 51.4% |
| Height at 11 years, mean (SD) z-score | -0.41 (1.03) | -0.25 (1.03) | -0.18 (1.01) | 0.29 (1.00) |
| Height at 16 years, mean (SD) z-score | -0.54 (1.03) | -0.35 (1.02) | -0.14 (1.16) | 0.27 (0.98) |
| **Confounding variables** |  |  |  |  |
| Social class |  |  |  |  |
| I Professional | 3.0% | 5.4% | 6.1% | 5.0% |
| II Intermediate | 14% | 18.2% | 23.6% | 43.1% |
| III Skilled Manual | 46.7% | 43.4% | 44.5% | 21.7% |
| III Skilled Non-Manual | 9.1% | 9.3% | 9.2% | 13.23% |
| IV Partly Skilled | 19.0% | 17.6% | 12.5% | 14.04% |
| V Unskilled | 8.2% | 6.1% | 4.1% | 2.94% |
| Maternal education, % post compulsory | 20.86% | 25.1% | 4.5% | 52.84% |
| Maternal height, mean (SD) cm | 160.81 (6.38) | 161.99 (6.45) | 161.28 (6.63) | 164.14 (6.93) |
| Paternal height, mean (SD) cm | 172.68 (8.07) | 174.47 (7.42) | 175.21 (7.51) | 178.28 (7.23) |

Note: Data are imputed (32 imputations); see methods for details and exact age of measurement in each cohort.

### Measures

#### Height

As described elsewhere,^23^ height was measured at the following ages in each study: ages 11 and 16 in the 1946c NSHD and 1958c NCDS; ages 10 and 16 in 1970c BCS; ages 7, 11, 14 and 17 in 2001c MCS. To account for age difference in exact age at measurement, and facilitate cross-cohort comparisons, in our main analysis, these were converted to age and sex-adjusted z-scores using the UK90 reference panel (the *childsds* package in R^24^). As a robustness check, we also converted height to age and cohort-specific z-scores, percentile ranks, and ridit scores.

#### Cognitive Function

Given the substantial heterogeneity in the cognitive tests administered to each cohort,^25^ we screened and identified measures which captured the same broad cognitive domains in childhood and adolescence. Assuming the relative ranking was similar, this would enable valid cross-cohort comparisons of associations with outcomes. Differences in the cognitive tests completed could potentially bias cross-cohort comparisons of cognition-height associations. Thus, we used multiple tests assessing different cognitive domains, measured at 2 different ages in order to examine whether our main findings were robust to the cognitive domain tested or age chosen.

##### Age 10/11 years

verbal reasoning test scores. These capture general verbal ability, verbal knowledge and reasoning. In the 1946c and 1958c this was measured using the verbal items from the General Ability Test (GAT).^26^ In the younger cohorts tests from the British Ability Scales (BAS), the Word Similarities test the precursor to the Verbal Similarities (BAS II) were administered in the 1970c and 2001c respectively.^27 28^

##### Age 14/16 years

reading/vocabulary test scores. The Watts-Vernon Reading Test (WVRT) was administered in the 1946c and the Reading Comprehension Test in the 1958c was devised by the National Foundation for Education Research (NFER) to parallel the aforementioned WVRT; both tests measure reading and word comprehension.^29^ The Applied Psychology Unit (APU) Vocabulary Test was administered in both the 1970c and 2001c, and measures word knowledge and meaning.^30^

Further, mathematic tests were administered in each cohort and used as an alternative indicator of cognitive performance. These were measured at 15/17 years in each cohort, at 7 years in 2001c and 10/11 years in the 1946c, 1958c and 1970c.

Since test performance differs by age,^25^ we constructed age-adjusted cognition measures by exporting the residuals from a linear regression model with test score as the outcome and age as the exposure variable (linear term). At age 16 years in 1970c, vocabulary tests were administered either in the home or in school—since test scores differed by mode, we also adjusted for a binary indicator of mode of administration in these models. Finally, to aid interpretation of effect sizes—and comparisons across age and cohort— in main analyses, we standardized all cognition measures to ridit scores (values between 0 and 1), with higher scores equating to better cognitive performance. In regression models coefficients thus show the mean difference in height comparing those with the lowest and highest cognition scores. Again, as a robustness check, we also repeated analyses converting cognition scores to (test and cohort-specific) z-scores and percentile ranks.

#### Potential Confounders

We hypothesized two sets of confounding variables—family socioeconomic characteristics, and parental height. Childhood socioeconomic position (SEP) was indicated by father’s occupational social class, measured at 10/11 years. To aid cross-cohort comparability, the Registrar General’s Social Class was used to classify social class—from I (professional), II (managerial and technical), IIIN (skilled nonmanual), IIIM (skilled manual), IV (partly-skilled), and V (unskilled) occupations. Maternal education was also used (ascertained at age 6 in 1946c NSHD, at birth in the 1958c NCDS and 1970c BCS, and at 9 months in 2001c MCS) with a binary indicator of whether the mother had left education at the mandatory leaving age (14 years old from 1918, 15 from 1944, and 16 from 1972).

Parental height was reported in early life—6 years (1946c), 11 years (1958c), 10 years (1970c), 9 months (2001c)—with those with particularly low height removed (height < 1.4m; n=14, 18, 12, 10 respectively in each cohort). Parental cognition was not measured in each cohort, precluding its investigation.

### Statistical Analysis

First, we conducted descriptive analyses and estimated bivariate correlations between cognition, social class, and height. Second, we regressed height on individual cognition variables (measured at the same age) adjusting for sex using linear regression. We did not anticipate childhood cognition and height to be directly causally related, but rather associated due to the influence of common environmental and genetic causes (see Supplementary Figure S1). To test our hypotheses, we sequentially adjusted for parental SEP (class and education) and parental height (mothers and fathers).

We next used quantile regression models^31^ to investigate whether mean differences observed in linear regression models arose due to differences in the lower quantiles (reflecting impaired growth, which may be more pronounced in older cohorts). We estimated the cognition-height association at each decile, again adjusting for sex and sequentially for parental SEP and parental height.

To address missing data, multiple imputation by chained equations was completed, with 32 imputations performed (burn in = 10 iterations). To improve the plausibility of the missing at random assumption, additional auxiliary variables were included imputation models (e.g., childhood BMI). OLS results were pooled using Rubin’s rules,^32^ while quantile regressions were pooled using the MI-then-bootstrap procedure^33^ with confidence intervals derived using the percentile method (200 bootstraps per imputation). Full details of the imputation procedure are provided in the Supplementary Information. The main findings did not differ when complete case analyses were conducted (results available upon request). Since the 1946c and 2001c are stratified study designs, analyses using these cohorts were conducted using study-specific design weights.

### Additional and sensitivity analyses

Since differences in missing data may bias cross-cohort patterns of association,^34^ in addition to using multiple imputation we carried out sensitivity analyses using pattern mixture modelling;^35^ our main OLS models were re-run altering imputed height values by a range of constant values (−1 to +1 SD).

We also carried out a series of other robustness checks. First, we repeated all models using different procedures for harmonizing height (raw, z-score, ridit, and rank score) and cognition variables (z-score and rank score). Second, we repeated analyses for males and females separately. Third, as the ethnic mix of the 2001c greatly exceeds that of older cohorts, we investigated if ethnic composition differences may have contributed to the main inferences drawn by conducting additional analyses restricted to the majority (European ancestry) population. Fourth, as height at age 16 was collected by self-report for some participants in the 1970c, we repeated main analyses using only the subset of those with measured height.

## Results

### Descriptive Statistics

The distributions of the cognition scores are shown in Figure 1 and sample descriptive statistics for height and covariates are shown in Table 1 (see also Supplementary Table S1 and Figure S1 for visual depictions). Childhood and adolescent height were greater in each successive cohort (Table 1). While differences in the tests administered precludes a comparison of absolute levels of cognition by cohort, the different tests used provided variation within each cohort and at each age (Figure 1). Correlations between the cognition scores and the height measures are shown in Figure 2 (correlation between all measures shown in Supplementary Figure S2). Cognitive test scores were strongly-moderately positively correlated with each other, with the size of the correlation weakening across time. Higher social class was associated with taller height and higher cognition scores; these correlations also appeared to weaken across time, as did positive correlations between maternal and paternal height (Supplementary Figure S3 and S4).

**Figure 1:**
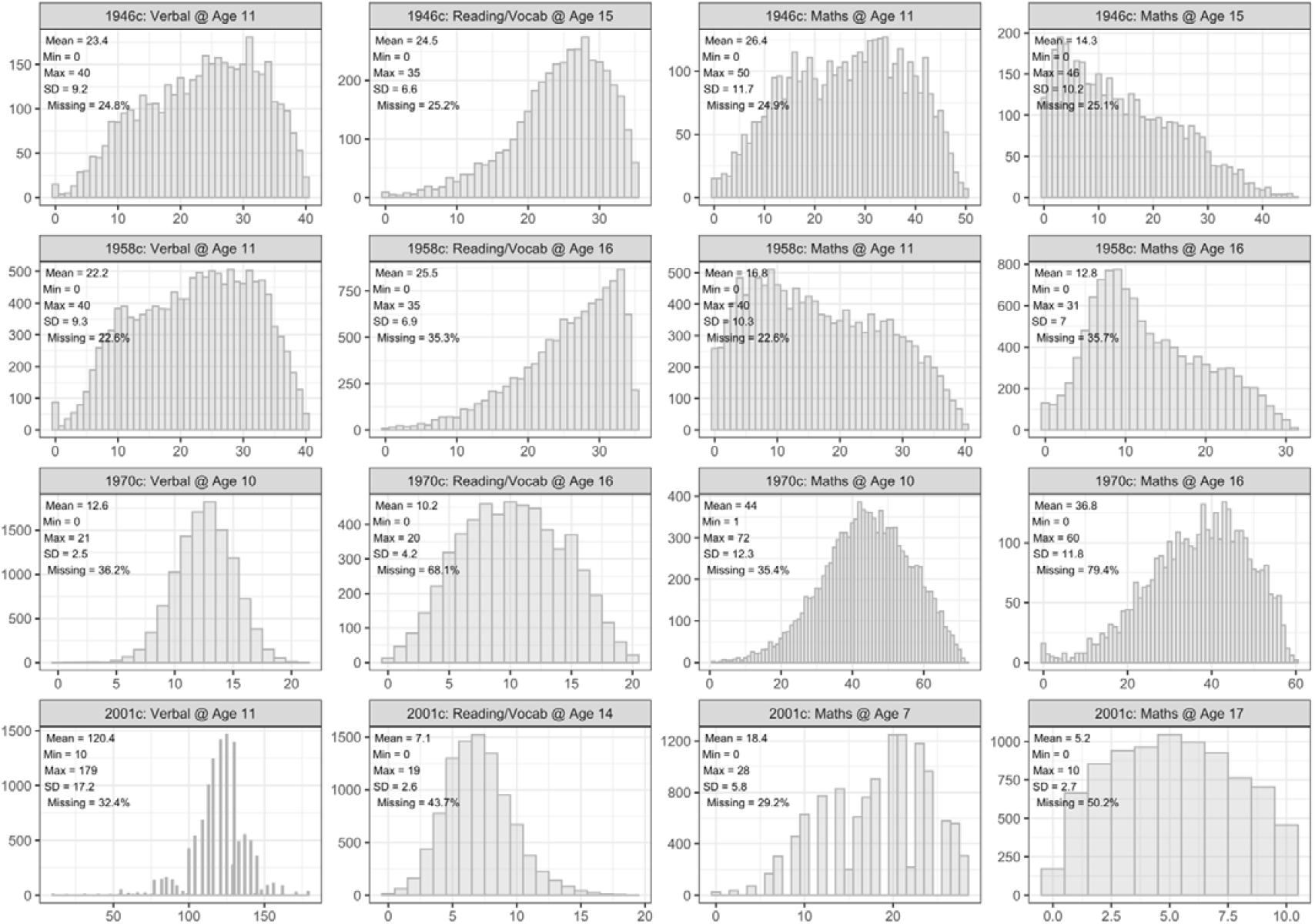
Distribution of raw cognition scores, by cohort and test. Observed data.

**Figure 2:**
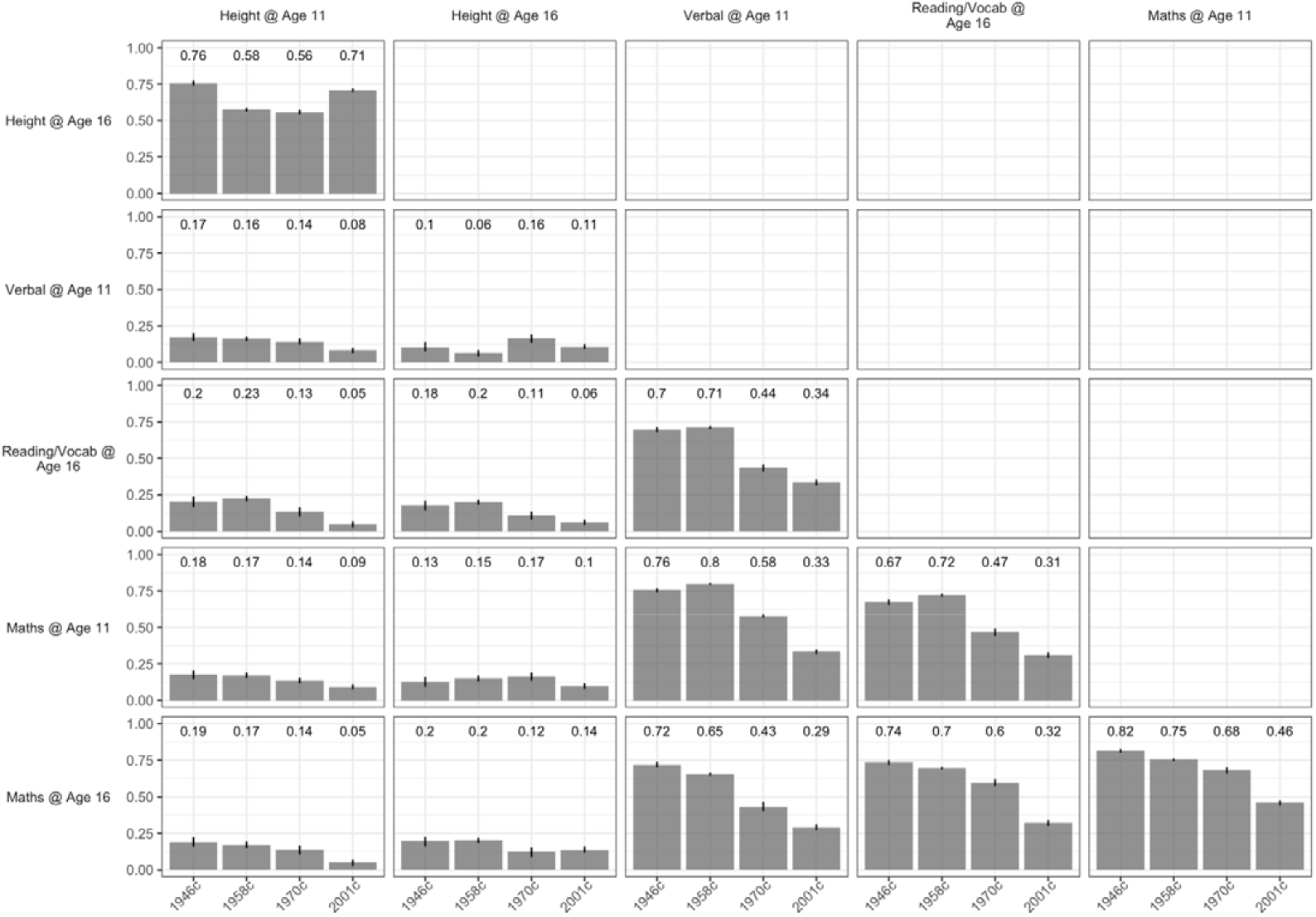
Spearman correlation between study variables by cohort. 95% CI intervals calculated using bootstrapping (200 bootstrap samples). Pairwise complete cases. Note, for correlations between cognition scores and height at the same stated age in the 2001c, contemporaneous height was used (e.g., as maths available for 7y only in 2001c, height at 7y was used in analyses for this correlation).

### Changing Association Between Cognition and Height

Higher cognition scores were associated with taller height—this association was found across all cohorts at both 11 and 16 years (Figure 3, left panel). These associations were stable or slightly larger between the 1946c and 1958c cohorts, and weakened thereafter, being substantially weakest in the youngest (2001c) cohort. For example, after adjustment for sex, the mean difference in height (Z-scores) comparing the highest with the lowest verbal reasoning score at 10/11 years was 0.56 SD (95% CI = 0.43, 0.7) in the 1946c, 0.58 SD (95% CI = 0.52, 0.64) in the 1958c, 0.46 SD (95% CI = 0.4, 0.53) in the 1970c, and 0.3 SD (95% CI = 0.23, 0.37) in the 2001c. Estimates from the 1946c were least precise owing to its smaller sample size, and confidence intervals overlapped with the larger 1958c. This pattern of weakening association across time was found at both 11 and 16 years, and across all measures of cognition (verbal reasoning, maths, and reading/vocabulary).

**Figure 3:**
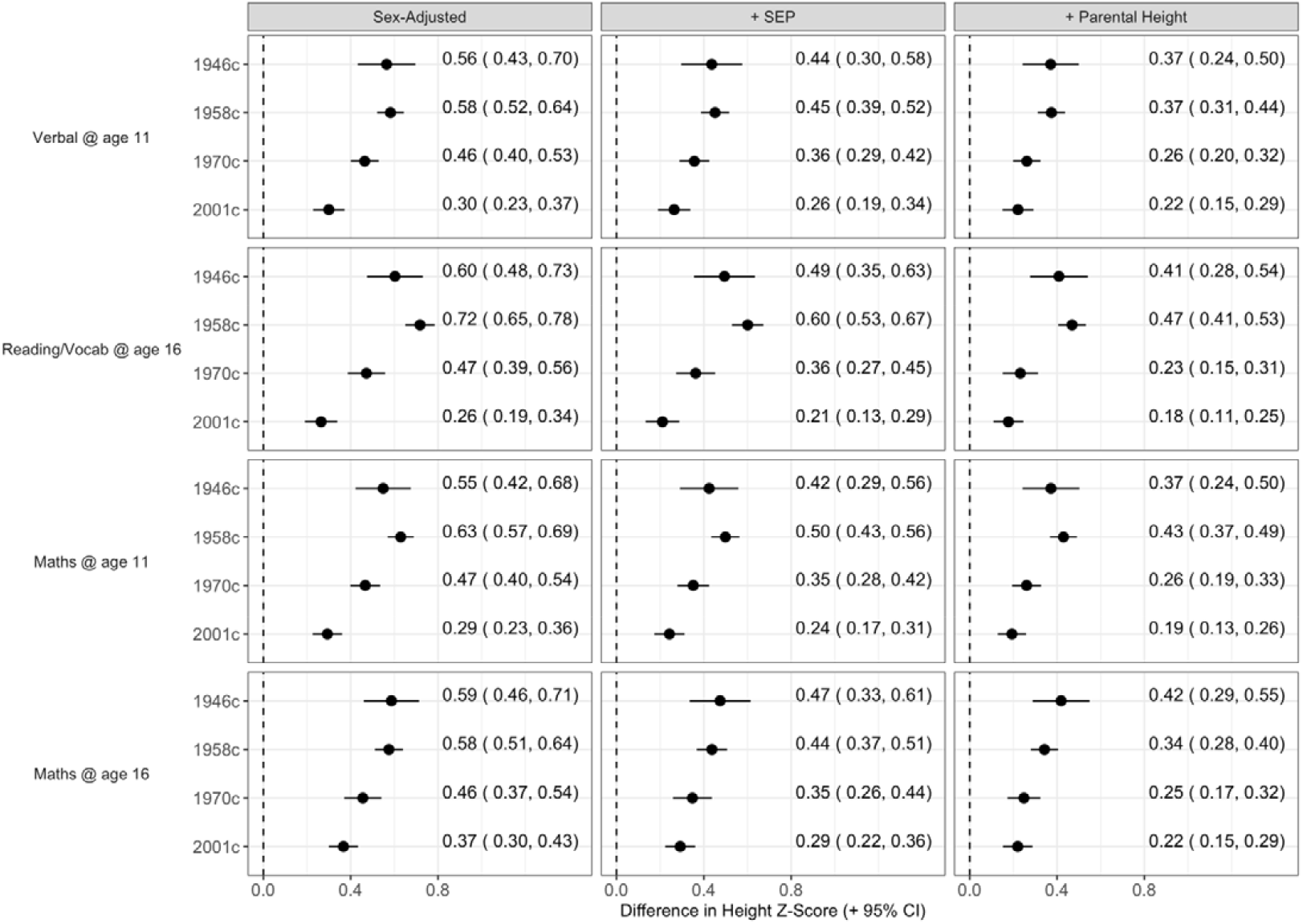
Association between height and cognition by cohort and test. Derived from pooled linear regression models (32 imputed datasets), adjusting for sex (left panel); sex, mother’s education and father’s social class (middle panel), and additionally for maternal and paternal height (right panel). Height converted to z-scores; cognition scores converted to ridit scores (range 0-1) – see methods. Estimates show mean differences in height comparing lowest versus highest cognition score.

Associations between cognition and height were partly attenuated after adjustment for parental SEP and partly attenuated further by parental height (Figure 3, middle and right panels, respectively). However, the associations did not attenuate completely to zero after this adjustment, and the weakening of the cognition-height association observed across each cohort was similar before and after adjustment. Patterns of association were also qualitatively similar within each sex (Supplementary Figure S5).

### Quantile Regressions

In quantile regression models adjusting for sex, there was evidence that associations between height and cognition were stronger at lower centiles of height, particularly in older cohorts (Figure 4). This was especially pronounced in the 1958c: for verbal scores at age 11, cognitive ability was associated with 0.73 SD (95% CI = 0.63, 0.82) greater height at the 10th percentile and 0.45 SD (95% CI = 0.34, 0.57) greater height at the 90th percentile. Differences across centile was less marked in the 2001c; corresponding figures in 2001c were 0.36 (95% CI = 0.24, 0.47) and 0.26 (95% CI = 0.14, 0.40), respectively. Qualitatively similar results were obtained when adjusted sequentially for childhood SEP (Supplementary Figure S6) and parental height (Supplementary Figure S7).

**Figure 4:**
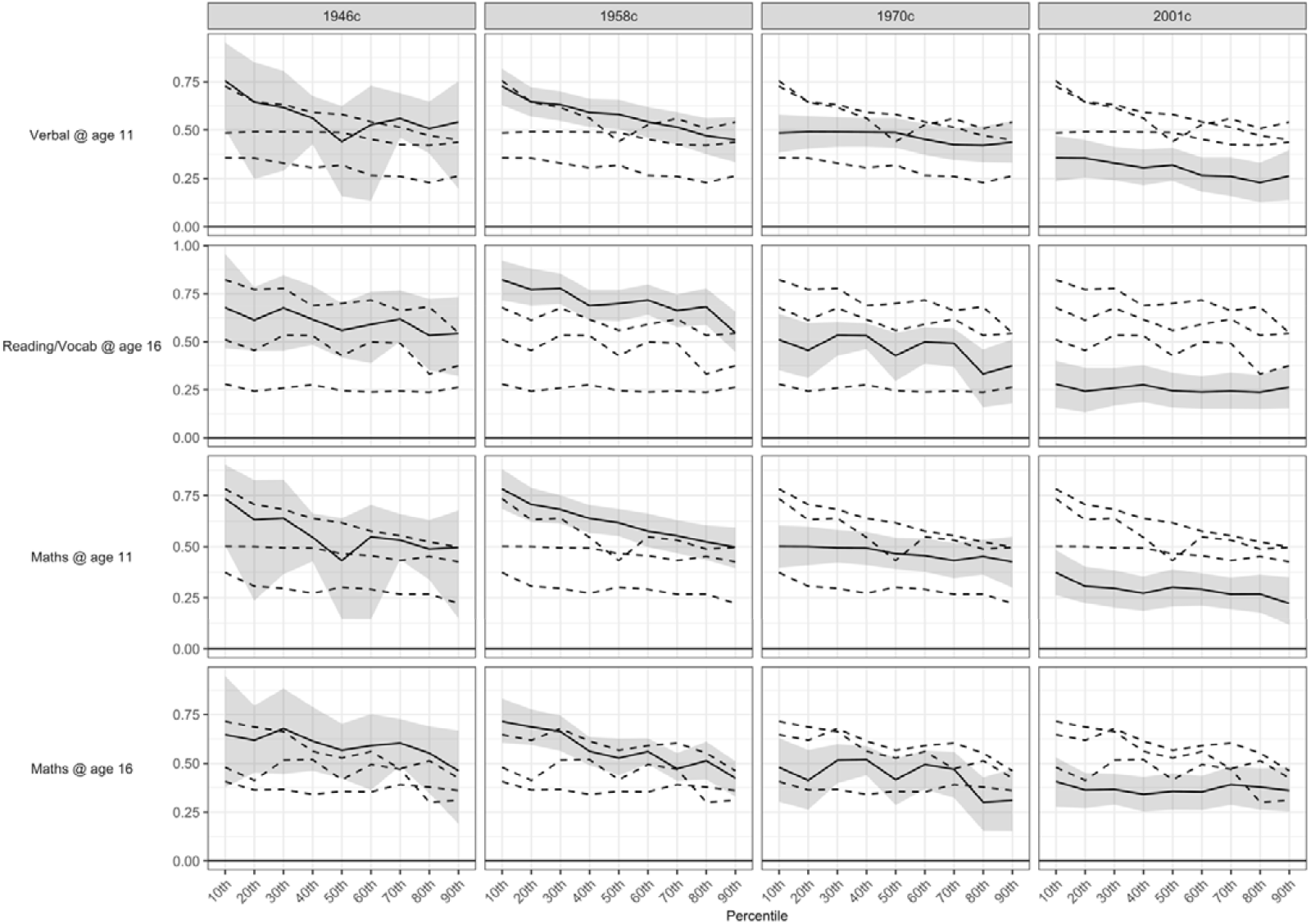
Association between height and cognition by cohort, test and height percentile. Derived from pooled quantile regression models (32 imputed datasets), adjusting for sex. Height covered to z-scores (see methods). Cognition scores converted to ridit scoring (range 0-1):estimates show the difference in height at each centile comparing lowest versus highest cognition score.

### Additional and Sensitivity Analyses

Among participants whose height was measured on at least one occasion, there were small differences in height according to whether an individual was lost to follow-up or not (< 0.3 SD). However, repeating main analyses altering imputed values by a constant factor to attempt to account for non-random missingness (i.e. pattern mixture modelling) generally yielded qualitatively similar results, even where the factor was large (i.e. > 0.8 SD; Supplementary Figures S8-S11): smallest associations were estimated in the 2001c, and largest associations were estimated in the 1946c or 1958c.

Repeating analyses using different procedures to score cognition (z-score, ridit scores and percentile ranks) and height (raw, z-score, rank, height charts, and ridit scoring) variables in order to compare estimates across cohorts also yielded qualitatively similar results to the main OLS analysis (Supplementary Figures S12-S14), as did limiting analyses with the 2001c to white participants only (Supplementary Figure S15), or the 1970c to those with measured rather than self-reported height at 16 years (Supplementary Figure S16).

## Discussion

### Summary of Findings

The association of higher cognition and height weakened substantially from 1957 to 2018. Associations were at least half as strong in the youngest (2001c) compared with the oldest (1946c and 1958c) cohorts. Quantile regression analyses suggested that height-cognition associations (and potentially their weakening across time) were driven by differences in lower quantiles of height. Associations were only partly attenuated when controlling for parental SEP or parental height.

Our findings build on earlier work, such as a study of Danish male conscripts,^15^ which also reported weakening of the cognition-height association from 1939 to 1967. We extend these findings by use of more recent data in more generalisable samples, utilizing multiple cognitive tests, undertaking both linear and quantile regression analyses, and accounting for both parental social class and height.

### Explanation of Findings

We anticipated two non-mutually exclusive processes which would cause associations between height and cognition. First, an association arising due to their shared genetic and environmental determinants. It is possible that changes in these—such as improvements in nutrition and reductions in the incidence (and/or impact of) infectious disease—led to a weakening of this association, particularly given the stronger magnitude of association at lower height centiles. This finding is consistent with declines in the socioeconomic patterning of height observed across this time period, as found in this study and reported elsewhere.^36^ We adjusted for multiple indicators of parental SEP available in each cohort (social class and education), and this may only partly capture the socioeconomically-graded environmental factors responsible for this association; further, we cannot fully rule out the potential for residual confounding by SEP itself (i.e. that height and cognition are correlated because of common effects of the familial environment). In particular, an apparent weakening of the correlation between parental social class and offspring cognition across time^37^ could have led to more residual confounding, and thus stronger associations between height and cognition in older cohorts. However, differences in the measurement of SEP across time may have operated in the other direction. While the SEP measures were constructed to be comparable in each cohort, the social class schema originated in the study of 20^th^ century job roles;^38^ analogously, parental education may have been a more distinguishing trait in earlier cohorts (before broader expansion of education). Thus, measurement error of SEP and resulting residual confounding may have instead been greater in the younger cohorts.

The robustness of the cross-cohort weakening in association after adjustment for parental height (an indicator of genetic liability to taller height) is similarly consistent with the hypothesis that environmental factors which influence early life height and cognition growth may be at least partly responsible for the weakening in association observed rather than directly inherited genetic factors related to height.

Second, it is also possible that changes in assortative mating could contribute to such associations; Keller et al. (2013) estimated that approximately half of the genetic association found in a single cohort was accounted for by assortative mating, and half by shared genetic factors.^3^ A weakening of assortative mating by cognition and height across cohorts could drive a weakening in the associations between these factors that we observed. However, we are unaware of empirical evidence investigating this trend, and are unable to directly test this owing to a lack of data for parental cognition. In the social science literature, evidence for changes in assortative mating by socioeconomic factors paints a complex picture: studies have suggested either stability,^39^ increases in assortative mating, or mixed findings.^40^ While we were unable to investigate changes in cognition-height assortative mating, parental height correlations were broadly similar in the 1946c, 1958c and 1970c and slightly weaker in the youngest cohort.

### Strengths and Limitations

Strengths of this study include the use of multiple national longitudinal cohorts from the UK—each with data on multiple cognitive test scores and measured height. We used four cohorts—the latest measure of the youngest cohort was 17 years—since height gains may occur up to 19-20 years,^41^ further work may be warranted in future to test if the patterns of association remain in later adulthood. Our analytical strategy is also a strength, enabling us to test the robustness of our findings across multiple ages and multiple domains of cognitive test performance and statistical methods.

Limitations include the potential for bias due to missing data, which is more pronounced in the youngest cohort. However, multiple imputation models were used, with multiple auxiliary variables included to increase the credibility of the ‘missing at random’ assumption such models entail. The multi-purpose and multidisciplinary cohorts used obtained short-form cognition tests—while these differed slightly in each cohort, we are not aware of a reason why this would lead to the pattern of result obtained. Inasmuch as this introduced random error, it would likely weaken observed associations due to regression dilution bias^42^ across all cohorts, and thus not bias cross-cohort comparisons. The broadly similar findings across the 4 distinct cognitive domains and at different ages suggest that the main findings were robust.

## Conclusions

Associations between higher cognition and taller height in childhood-adolescence weakened markedly from 1957 to 2018. These findings support the notion that changes to environmental and societal factors across time may substantially weaken associations between cognition and other traits.

## Data Availability

Data files are available online via the UK Data Service (1958c, 1970c, and 2001c; https://www.data-archive.ac.uk/) and via application (1946c; https://skylark.ucl.ac.uk/).

https://skylark.ucl.ac.uk/

## Acknowledgements

We thank Rebecca Hardy for helpful comments on earlier draft results.

## Author contributions

DB, VM, LW, NMD conceived and designed the study. LW analysed all the data presented in this paper and independently replicated the findings from earlier analyses conducted by DB. DB wrote the first draft. All authors provided critical revisions and all authors read and approved the submitted manuscript.

## Data availability

Data files are available through the UK Data Service (1958c, 1970c, and 2001c; https://www.data-archive.ac.uk/) and via application (1946c; https://skylark.ucl.ac.uk/). The code used to run the analysis is available at https://osf.io/54unw/.

## References

1. Deary IJ, Batty GD. Cognitive epidemiology. J Epidemiol Community Health 2007;61(5):378–84.

2. Marioni RE, Batty GD, Hayward C, et al. Common genetic variants explain the majority of the correlation between height and intelligence: the generation Scotland study. Behav Genet 2014;44(2):91–96.

3. Keller MC, Garver-Apgar CE, Wright MJ, et al. The genetic correlation between height and IQ: Shared genes or assortative mating? PLoS genetics 2013;9(4):e1003451.

4. Monteiro POA, Victora CG. Rapid growth in infancy and childhood and obesity in later life - a systematic review. Obes Rev 2005;6(2):143–54.

5. Walker SP, Wachs TD, Gardner JM, et al. Child development: risk factors for adverse outcomes in developing countries. Lancet 2007;369(9556):145–57.

6. Perkins JM, Subramanian SV, Davey Smith G, et al. Adult height, nutrition, and population health. Nutr Rev 2016;74(3):149–65.

7. Batty GD, Shipley MJ, Gunnell D, et al. Height, wealth, and health: An overview with new data from three longitudinal studies. Econ Hum Biol 2009;7(2):137–52. doi: doi:DOI:10.1016/j.ehb.2009.06.004

8. Silventoinen K, Iacono WG, Krueger R, et al. Genetic and environmental contributions to the association between anthropometric measures and IQ: a study of Minnesota twins at age 11 and 17. Behav Genet 2012;42(3):393–401.

9. Vuoksimaa E, Panizzon MS, Franz CE, et al. Brain structure mediates the association between height and cognitive ability. Brain Structure and Function 2018;223(7):3487–94.

10. Jelenkovic A, Sund R, Hur Y-M, et al. Genetic and environmental influences on height from infancy to early adulthood: An individual-based pooled analysis of 45 twin cohorts. Sci Rep 2016;6(1):1–13.

11. Meredith HV. Findings from Asia, Australia, Europe, and North America on secular change in mean height of children, youths, and young adults. Am J Phys Anthropol 1976;44(2):315–25.

12. Cole TJ. Secular trends in growth. Proc Nutr Soc 2000;59(02):317–24.

13. Bratsberg B, Rogeberg O. Flynn effect and its reversal are both environmentally caused. Proc Natl Acad Sci USA 2018;115(26):6674–78.

14. Gale C. Commentary: height and intelligence. Int J Epidemiol 2005;34(3):678–79.

15. Teasdale TW, Sørensen T, Owen DR. Fall in association of height with intelligence and educational level. BMJ: British Medical Journal 1989;298(6683):1292.

16. Wadsworth M, Kuh D, Richards M, et al. Cohort profile: The 1946 National Birth Cohort (MRC National Survey of Health and Development). Int J Epidemiol 2006;35(1):49–54.

17. Power C, Elliott J. Cohort profile: 1958 British birth cohort (National Child Development Study). Int J Epidemiol 2006;35(1):34–41.

18. Elliott J, Shepherd P. Cohort profile: 1970 British birth cohort (BCS70). Int J Epidemiol 2006;35(4):836–43.

19. Connelly R, Platt L. Cohort Profile: UK Millennium Cohort Study (MCS). Int J Epidemiol 2014;43(6):1719–25.

20. Shepherd P, Gilbert E. Millenium Cohort Study ethical review and consent, 2019.

21. Centre for Longitudinal Studies. National child development study ethical review and consent, 2014.

22. Shepherd P, Gilbert E. British Cohort Study ethical review and consent, 2019.

23. Johnson W, Li L, Kuh D, et al. How Has the Age-Related Process of Overweight or Obesity Development Changed over Timeã Co-ordinated Analyses of Individual Participant Data from Five United Kingdom Birth Cohorts. PLoS Med 2015;12(5):e1001828.

24. Vogel M. A Combined Approach to Generate Age & Sex Dependent Reference Intervals in Pediatrics. QUCOSA: Diss, 2018.

25. Moulton V, McElroy E, Richards M, et al. A guide to the cognitive measures in the British birth cohort studies. CLOSER, UK 2019

26. Pigeon D. Tests used in the 1954 and 1957 surveys In Douglas JWB, editor.(Ed.), The home and the school: A study of ability and attainment in the primary school (pp. 129–132). London, England: Macgibbon and Kee[Google Scholar] 1964

27. Elliot C, Murray D, Pearson L. British Ability Scales. Windsor, UK, 1978.

28. CD E, P S, K M. British Ability Scales Second Edition (BAS II) Administration and Scoring Manual. London, 1996.

29. Shepherd P. 1958 National Child Development Study user guide: Measures of ability at ages 7 to 16: University of London, Institute of Education, Centre for Longitudinal…, 2012.

30. Closs S. APU Vocabulary Test (multiple choice format). Kent, 1986.

31. Koenker R, Bassett Jr G. Regression quantiles. Econometrica 1978:33–50.

32. Rubin DB. Inference and missing data. Biometrika 1976;63(3):581–92.

33. Bartlett JW, Hughes RA. Bootstrap inference for multiple imputation under uncongeniality and misspecification. Stat Methods Med Res 2020;29(12):3533–46.

34. Bann D, Wright L, Goisis A, et al. Investigating change across time in prevalence or association using observational data: guidance on utility, methodology, and interpretation. OSF Preprints 2021

35. Leurent B, Gomes M, Faria R, et al. Sensitivity analysis for not-at-random missing data in trial-based cost-effectiveness analysis: a tutorial. Pharmacoeconomics 2018;36(8):889–901.

36. Bann D, Johnson W, Li L, et al. Socioeconomic inequalities in childhood and adolescent body-mass index, weight, and height from 1953 to 2015: an analysis of four longitudinal, observational, British birth cohort studies. The Lancet Public Health 2018

37. Paterson L. Filial intelligence and family social class, 1947 to 2012. Sociological Science 2021;8:325–45.

38. Bland R. Measuring” Social Class” A Discussion of the Registrar-General’s Classification. Sociology 1979;13(2):283–91.

39. Henz U, Mills C. Social class origin and assortative mating in Britain, 1949–2010. Sociology 2018;52(6):1217–36.

40. Schwartz CR. Trends and variation in assortative mating: Causes and consequences. Annu Rev Sociol 2013;39:451–70.

41. Rodriguez-Martinez A, Zhou B, Sophiea MK, et al. Height and body-mass index trajectories of school-aged children and adolescents from 1985 to 2019 in 200 countries and territories: a pooled analysis of 2181 population-based studies with 65 million participants. Lancet 2020;396(10261):1511–24.

42. Hutcheon JA, Chiolero A, Hanley JA. Random measurement error and regression dilution bias. BMJ 2010;340:c2289.

