## supplemental file for "Weakening of the cognition and height association from 1957 to 2018: findings from four British birth cohort studies"

Supplementary Information

### Multiple Imputation Procedure

We use multiple imputation by chained equations to impute missing data. Our sample in our imputation models was all individuals with observed sex data who were singleton births from England, Scotland, or Wales. We performed imputation for each cohort/sex combination separately. Continuous variables were imputed using the random forest algorithm, binary variables using logistic regression models and other categorical variables using multinomial logistic regression models. We created 32 imputed datasets per cohort/sex, using a burn-in of 10 iterations per imputed dataset.

All variables used in the final analysis were included in the imputation models, including survey weights, which were included as continuous terms. Participant height variables were added as raw scores, with age of measurement included as imputation covariates. Cognition data was inputted as raw residuals. The reason for these choices was that we needed to convert the variables into other scores (ridit, Z-scores, ranks, etc.) post imputation. We also included a set of auxiliary variables to increase imputation accuracy: BMI at each age of height measurement and father’s education level (compulsory education or lower vs higher than compulsory education) for each cohort; and for the 1970c, reading comprehension and whether height was self-reported, each measured at age 16.

### Tables

Table S1: Descriptive statistics, observed and imputed data. Imputed data drawn from 32 imputations.

|  | | 1946c | | | 1958c | | | 1970c | | | 2001c | | |
| --- | --- | --- | --- | --- | --- | --- | --- | --- | --- | --- | --- | --- | --- |
|  | Variable | Observed | Missing % | Imputed | Observed | Missing % | Imputed | Observed | Missing % | Imputed | Observed | Missing % | Imputed |
|  | Sample Size | 5,348 |  | 5,348 | 17,490 |  | 17,490 | 17,617 |  | 17,617 | 17,057 |  | 17,057 |
| Sex | Female | 2,538 (47.46%) | 0% | 2,535.5 (47.41%) | 8,445 (48.28%) | 0% | 8,445 (48.28%) | 8,468 (48.07%) | 0% | 8,468 (48.07%) | 8,275 (48.51%) | 0% | 8,287.61 (48.59%) |
|  | Male | 2,810 (52.54%) |  | 2,812.5 (52.59%) | 9,045 (51.72%) |  | 9,045 (51.72%) | 9,149 (51.93%) |  | 9,149 (51.93%) | 8,782 (51.49%) |  | 8,769.39 (51.41%) |
|  | Height @ Age 11 (Z-Score) | -0.33 (1.04) | 26.61% | -0.41 (1.03) | -0.22 (1.02) | 30.51% | -0.25 (1.03) | -0.17 (1.01) | 30.45% | -0.18 (1.01) | 0.27 (1.03) | 31.88% | 0.29 (1) |
|  | Height @ Age 16 (Z-Score) | -0.47 (1.04) | 32.24% | -0.54 (1.03) | -0.33 (1.02) | 40.21% | -0.35 (1.02) | -0.09 (1.18) | 58.36% | -0.14 (1.16) | 0.23 (1.01) | 41.12% | 0.27 (0.98) |
|  | Maternal Height (cm) | 161.2 (6.35) | 21.35% | 160.81 (6.38) | 162.02 (6.46) | 26.23% | 161.99 (6.45) | 161.3 (6.65) | 25.58% | 161.28 (6.63) | 163.65 (7.03) | 5.83% | 164.14 (6.93) |
|  | Paternal Height (cm) | 173.4 (7.94) | 35.71% | 172.68 (8.07) | 174.53 (7.42) | 27.94% | 174.47 (7.42) | 175.24 (7.53) | 28.91% | 175.21 (7.51) | 177.85 (7.41) | 31.45% | 178.28 (7.23) |
| Father's Social Class | I Professional | 238 (6.08%) | 26.83% | 161.31 (3.02%) | 689 (5.56%) | 29.13% | 952.12 (5.44%) | 751 (6.27%) | 32.05% | 1,078.22 (6.12%) | 469 (5.17%) | 46.8% | 850.12 (4.98%) |
|  | II Intermediate | 761 (19.45%) |  | 748.82 (14%) | 2,290 (18.47%) |  | 3,182.47 (18.2%) | 2,867 (23.95%) |  | 4,160.09 (23.61%) | 3,834 (42.25%) |  | 7,354.00 (43.11%) |
|  | III Skilled Manual | 1,331 (34.01%) |  | 2,498.96 (46.73%) | 5,359 (43.23%) |  | 7,585.34 (43.37%) | 5,287 (44.17%) |  | 7,833.41 (44.47%) | 2,030 (22.37%) |  | 3,700.60 (21.7%) |
|  | III Skilled Non-Manual | 601 (15.36%) |  | 484.02 (9.05%) | 1,166 (9.41%) |  | 1,622.09 (9.27%) | 1,105 (9.23%) |  | 1,621.81 (9.21%) | 1,197 (13.19%) |  | 2,257.30 (13.23%) |
|  | IV Partly Skilled | 742 (18.96%) |  | 1,015.55 (18.99%) | 2,152 (17.36%) |  | 3,076.56 (17.59%) | 1,478 (12.35%) |  | 2,198.56 (12.48%) | 1,288 (14.19%) |  | 2,394.23 (14.04%) |
|  | V Unskilled | 240 (6.13%) |  | 439.34 (8.22%) | 740 (5.97%) |  | 1,071.41 (6.13%) | 483 (4.03%) |  | 724.91 (4.11%) | 256 (2.82%) |  | 500.75 (2.94%) |
| Mother's Education | High | 1,235 (28.54%) | 19.07% | 1,115.51 (20.86%) | 4,224 (24.93%) | 3.14% | 4,384.25 (25.07%) | 5,681 (34.44%) | 6.37% | 6,069.03 (34.45%) | 8,472 (49.86%) | 0.39% | 9,013.27 (52.84%) |
|  | Low | 3,093 (71.46%) |  | 4,232.49 (79.14%) | 12,717 (75.07%) |  | 13,105.75 (74.93%) | 10,813 (65.56%) |  | 11,547.97 (65.55%) | 8,518 (50.14%) |  | 8,043.73 (47.16%) |

### Figures

a)	Common causes	b) Assortative mating

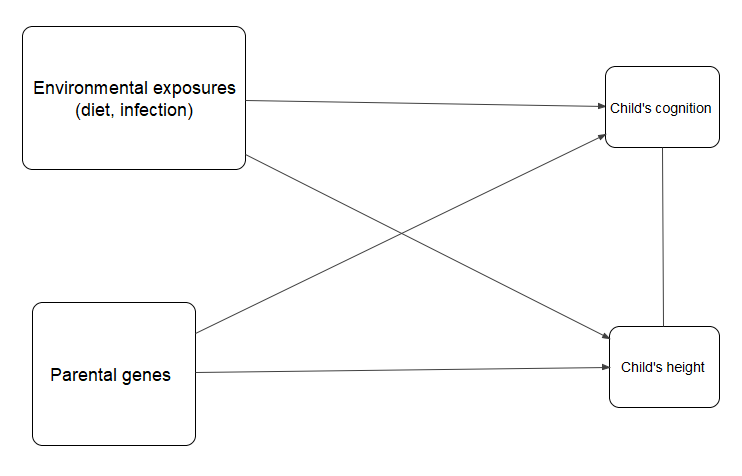

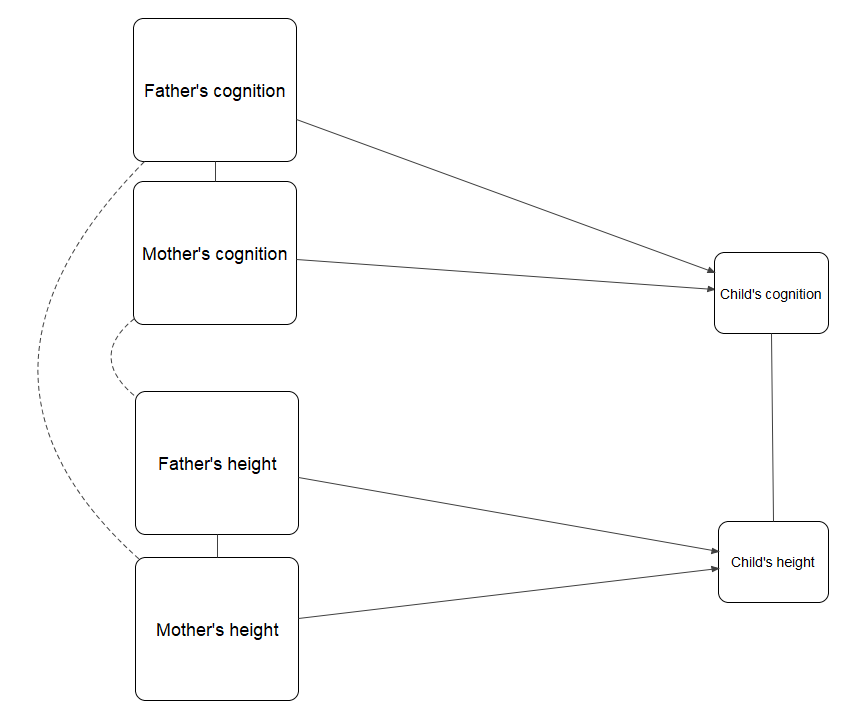

*Figure S1. An illustrative causal diagram (Directed Acyclic Graph) of two alternative processes which may either (or both in some combination) generate associations between child cognition and height: a) shared genetic and environmental factors and b) assortative mating of parental cognition and height (within and across each trait).*

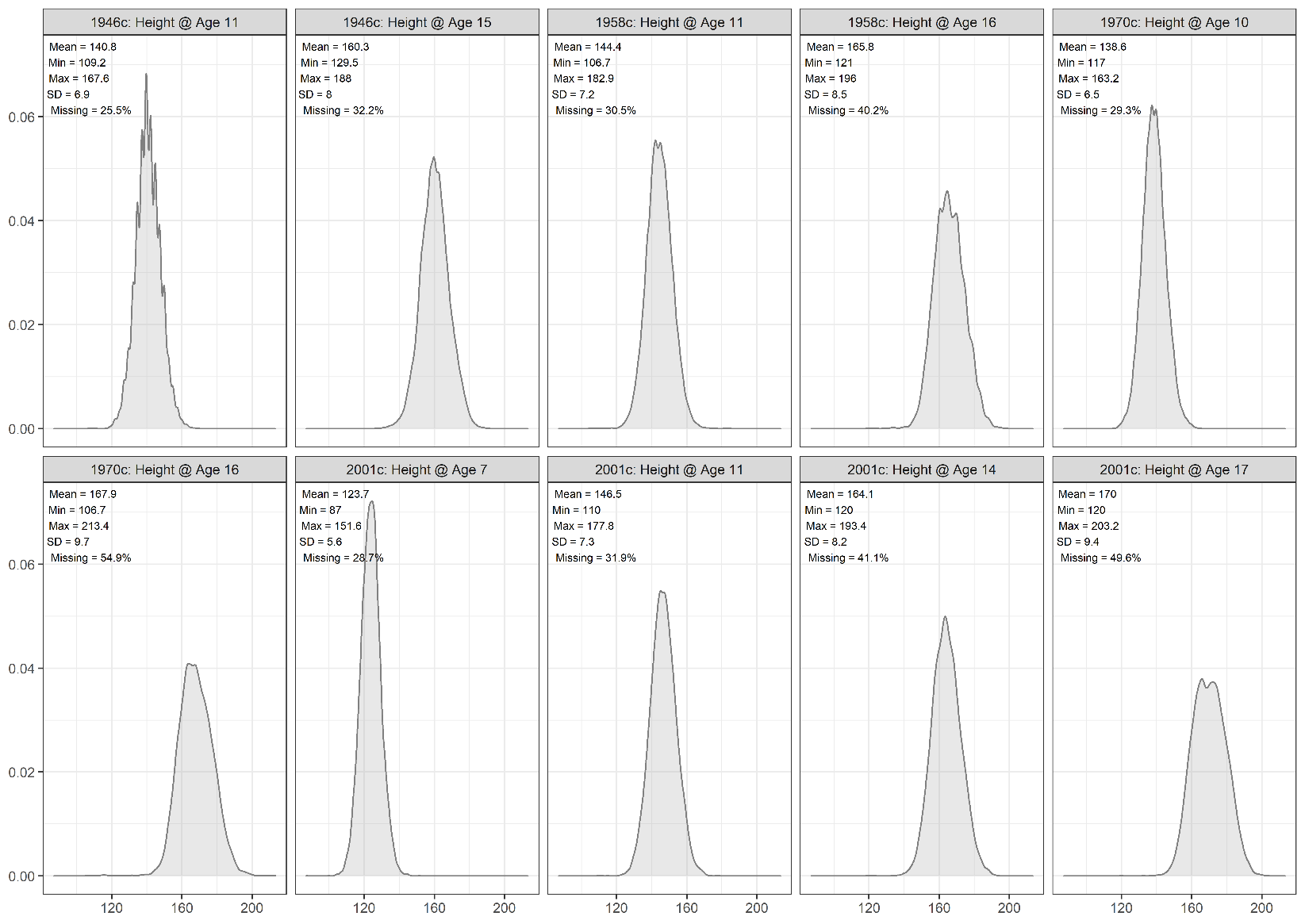

Figure S2: Distribution of height by cohort and age, observed data.

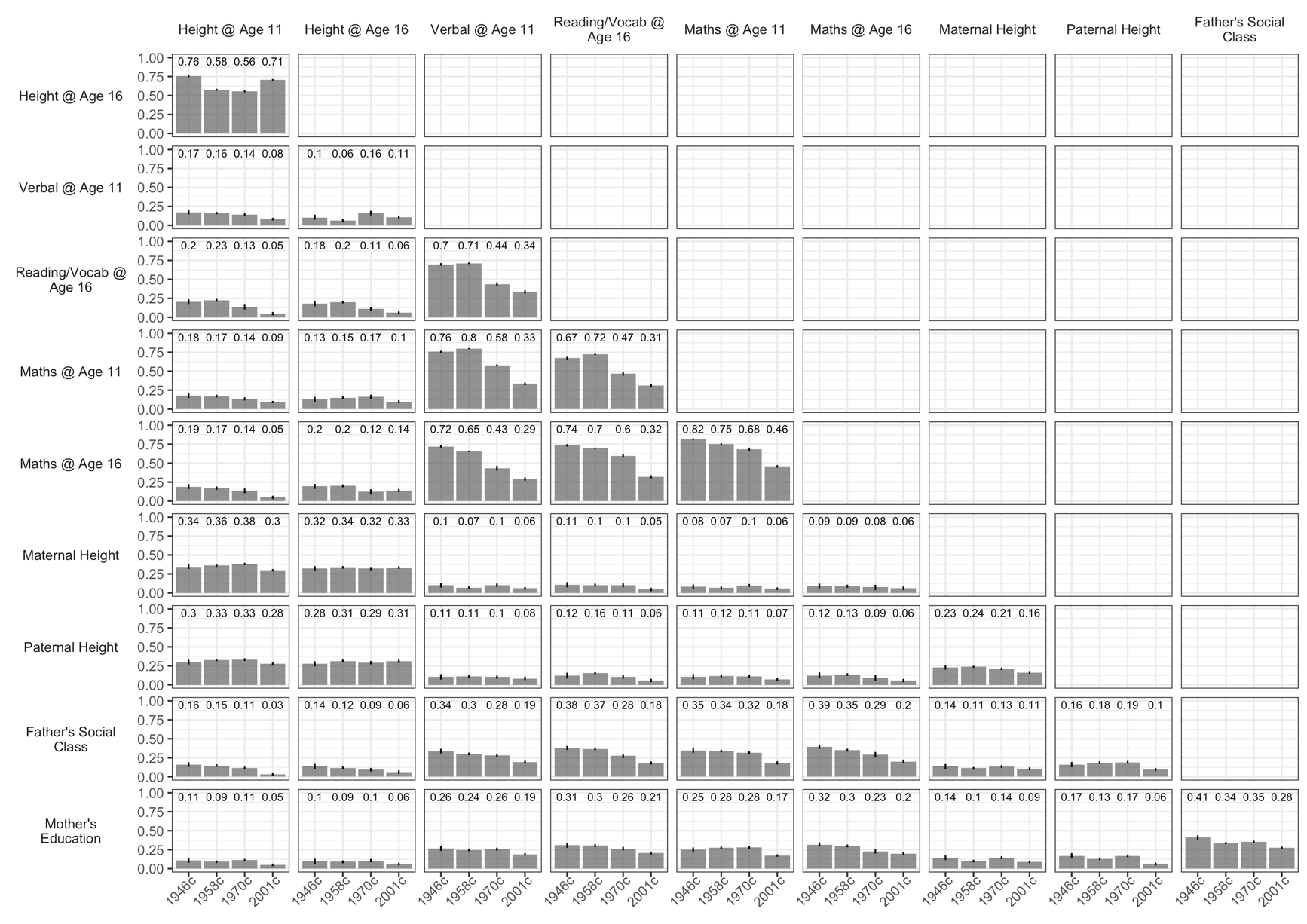

Figure S3: Spearman correlation between study variables by cohort. 95% CI intervals calculated using bootstrapping (200 bootstrap samples). Observed data.

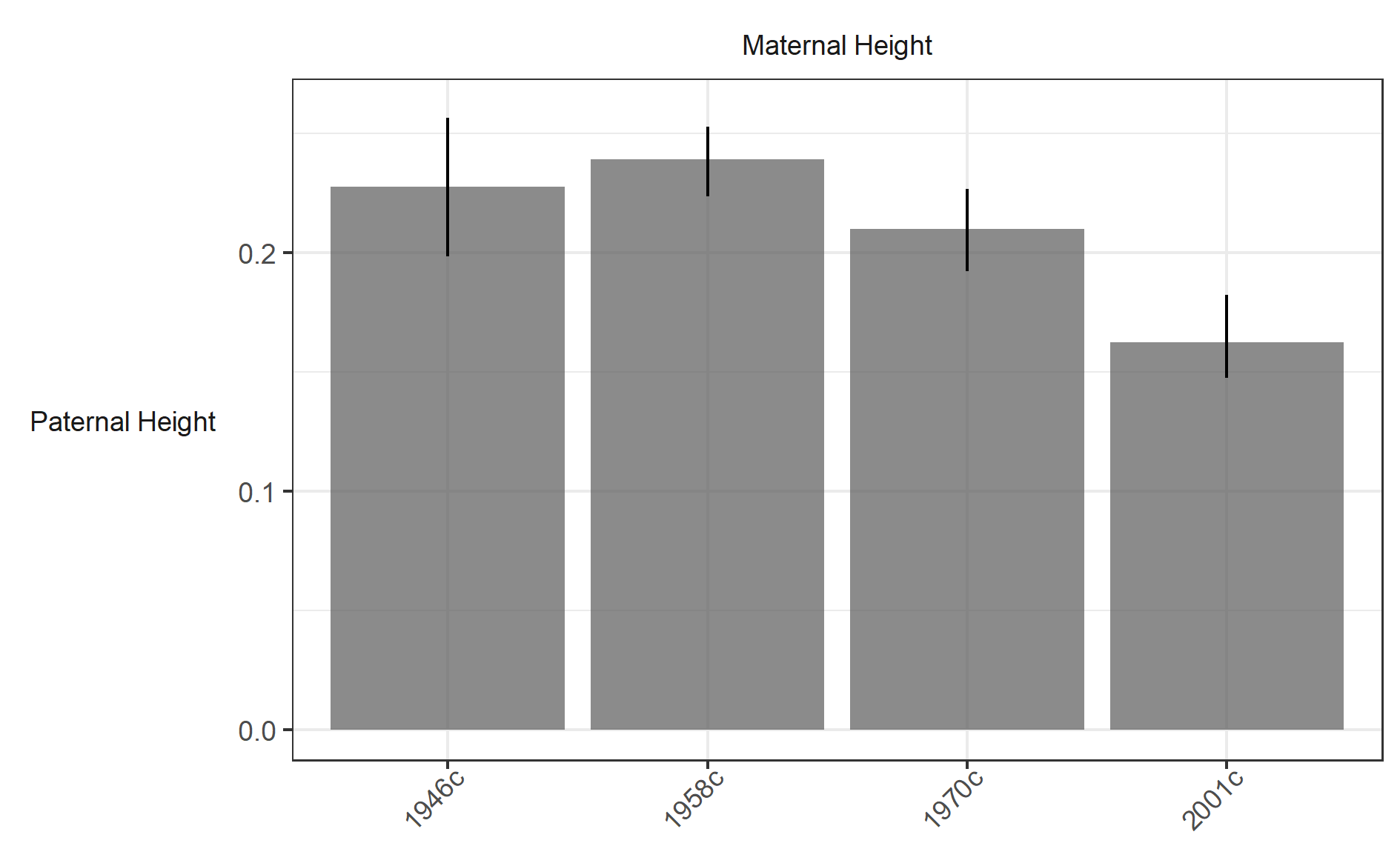

Figure S4: Spearman correlation between paternal and maternal height by cohort. 95% CI intervals calculated using bootstrapping (200 bootstrap samples).

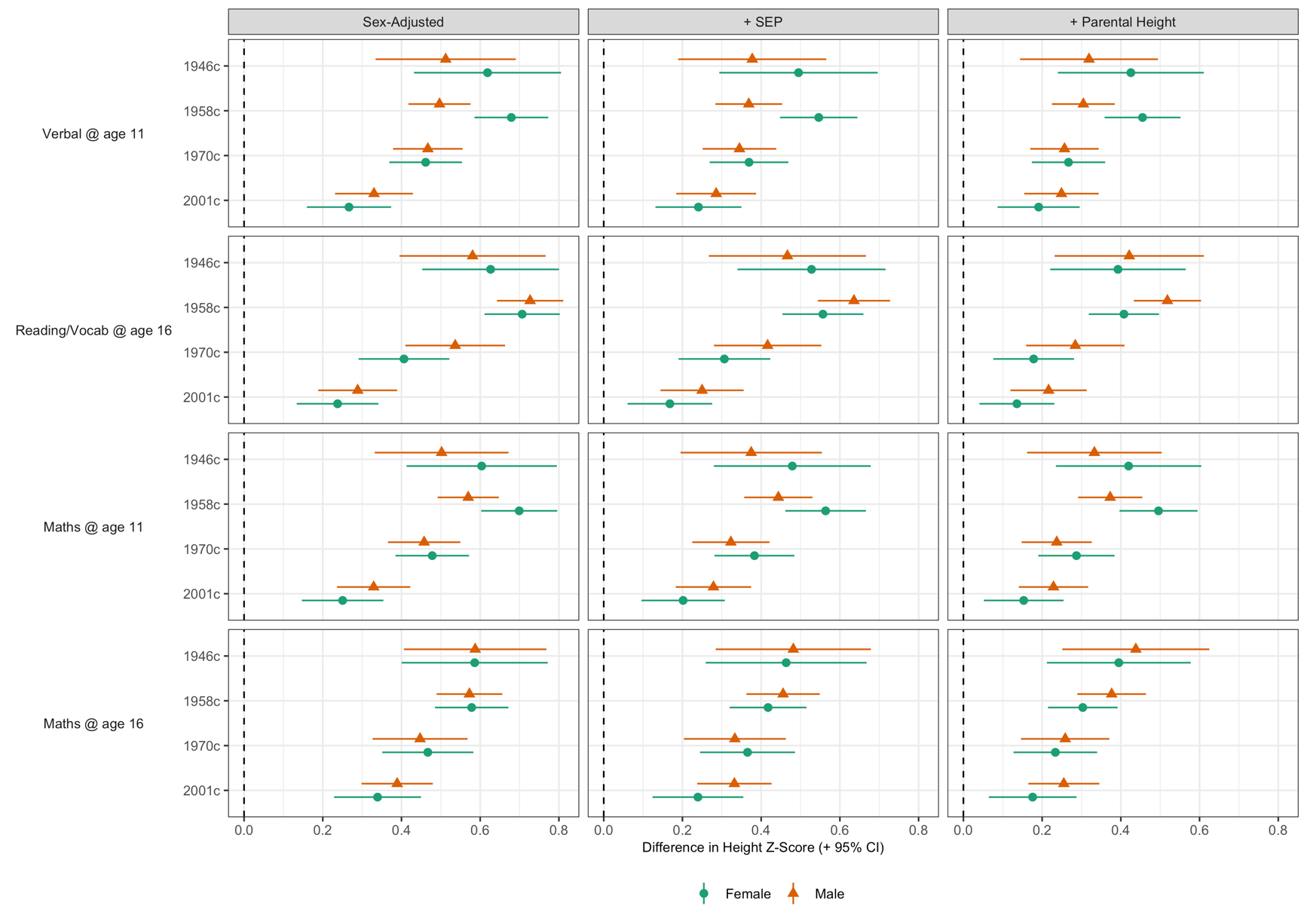

Figure S5: Association between height and cognition by cohort, test and sex. Derived from pooled OLS models (32 imputed datasets), including no other covariates (left panel); adjusting for mother’s education and father’s social class (middle panel); and adjusting for mother’s education, father’s social class, and paternal and maternal height (right panel). Height harmonised across cohorts using growth charts from 1990 UK growth study. Cognition scores harmonised using ridit scoring (range 0-1).

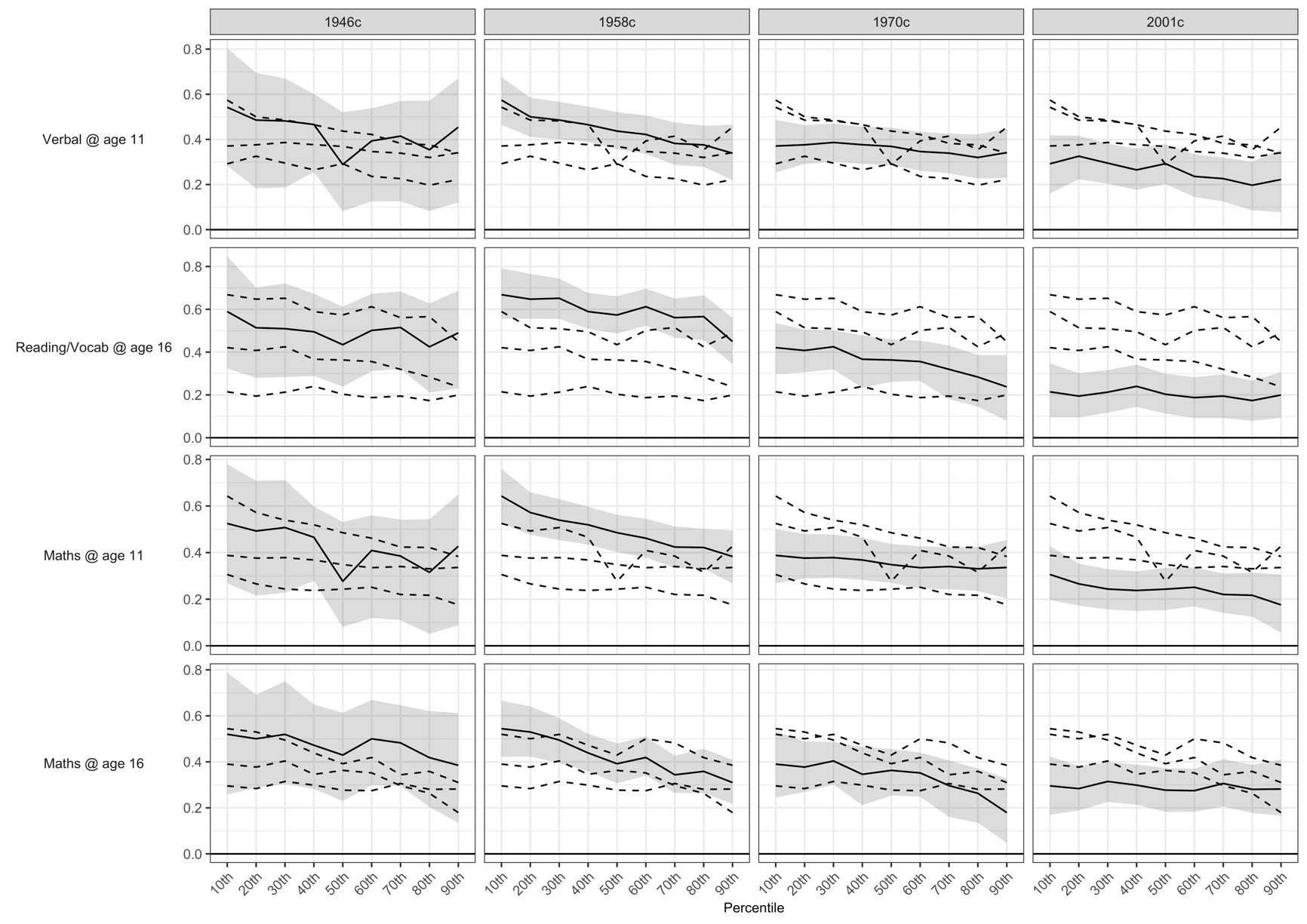

Figure S6: Association between height and cognition by cohort, test and height percentile. Derived from pooled quantile regression models (32 imputed datasets), adjusting for sex, mother’s education and father’s social class. Height harmonised across cohorts using growth charts from 1990 UK growth study. Cognition scores harmonised using ridit scoring (range 0-1).

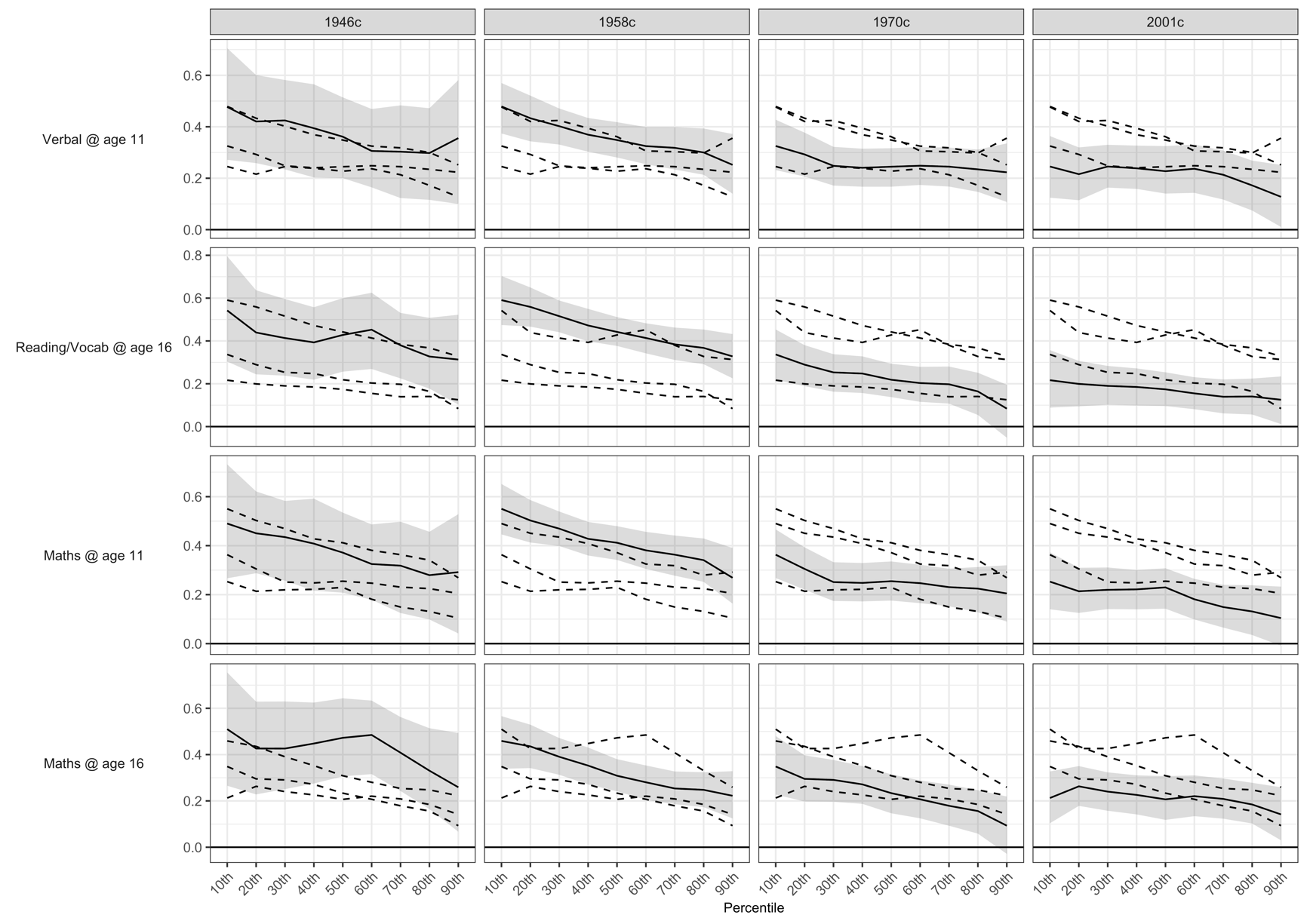

Figure S7: Association between height and cognition by cohort, test and height percentile. Derived from pooled quantile regression models (32 imputed datasets), adjusting for sex, mother’s education, father’s social class and maternal and paternal height. Height harmonised across cohorts using growth charts from 1990 UK growth study. Cognition scores harmonised using ridit scoring (range 0-1).

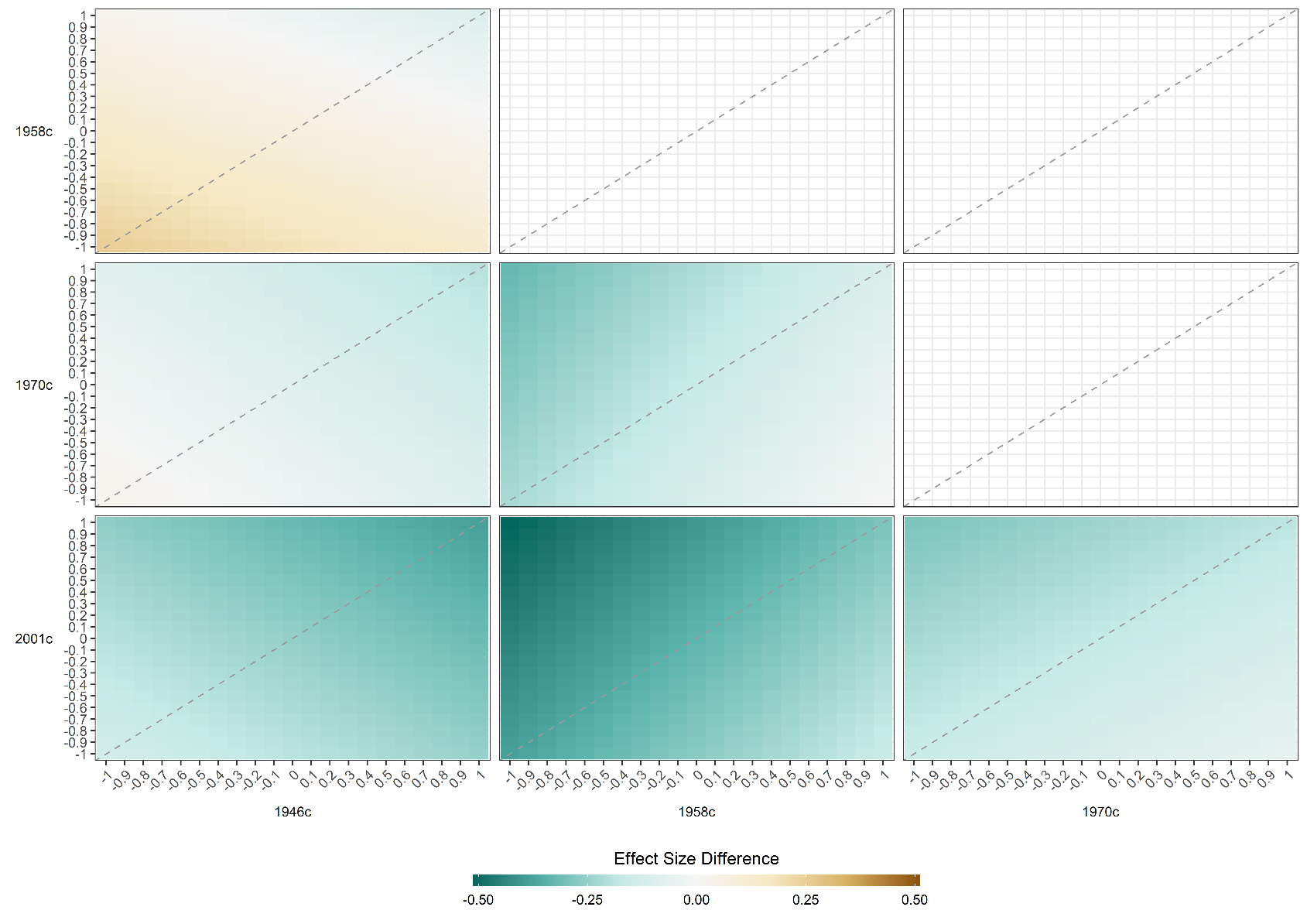

Figure S8: Result of pattern mixture models. Difference in association between height and maths @ age 11 across cohort. Derived from pooled OLS models (32 imputed datasets) including adjustment for sex. OLS models repeated with imputed height values adjusted by a constant factor given in columns and rows. The colour of each square indicates difference in the association between height and cognition between two cohorts with the coefficient for the cohort on the horizontal subtracted from that on the horizontal (with given adjustment factors used). Height harmonised across cohorts using growth charts from 1990 UK growth study. Cognition scores harmonised using ridit scoring (range 0-1).

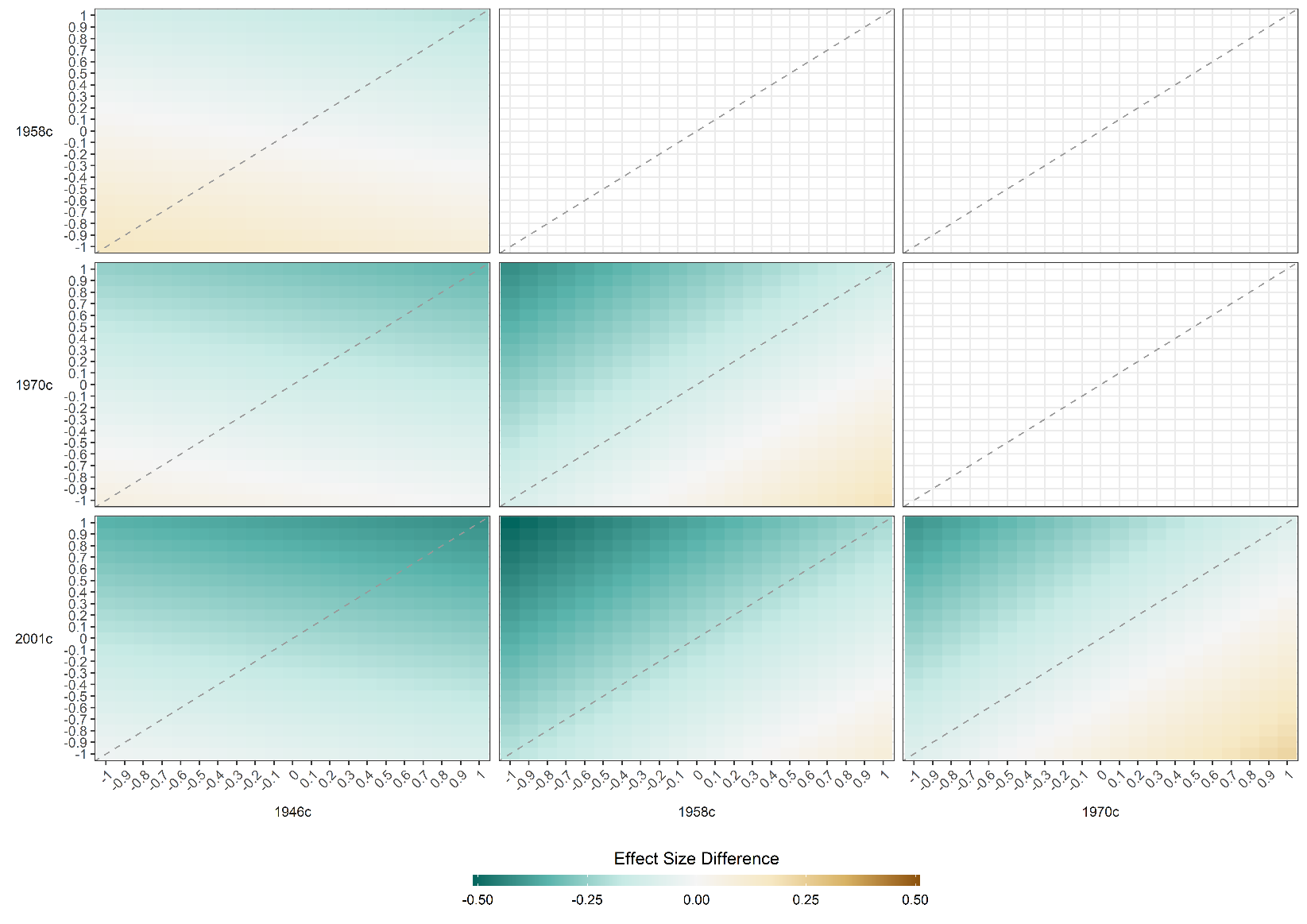

Figure S9: Result of pattern mixture models. Difference in association between height and maths @ age 16 across cohort. Derived from pooled OLS models (32 imputed datasets) including adjustment for sex. OLS models repeated with imputed height values adjusted by a constant factor given in columns and rows. The colour of each square indicates difference in the association between height and cognition between two cohorts with the coefficient for the cohort on the horizontal subtracted from that on the horizontal (with given adjustment factors used). For instance, in the bottom left panel, the co-ordinate 1, -1 refers to difference in coefficients where +1 SD was added to imputed height values in 1946c and -1 SD was added to imputed height values in 2001c. Height harmonised across cohorts using growth charts from 1990 UK growth study. Cognition scores harmonised using ridit scoring (range 0-1).

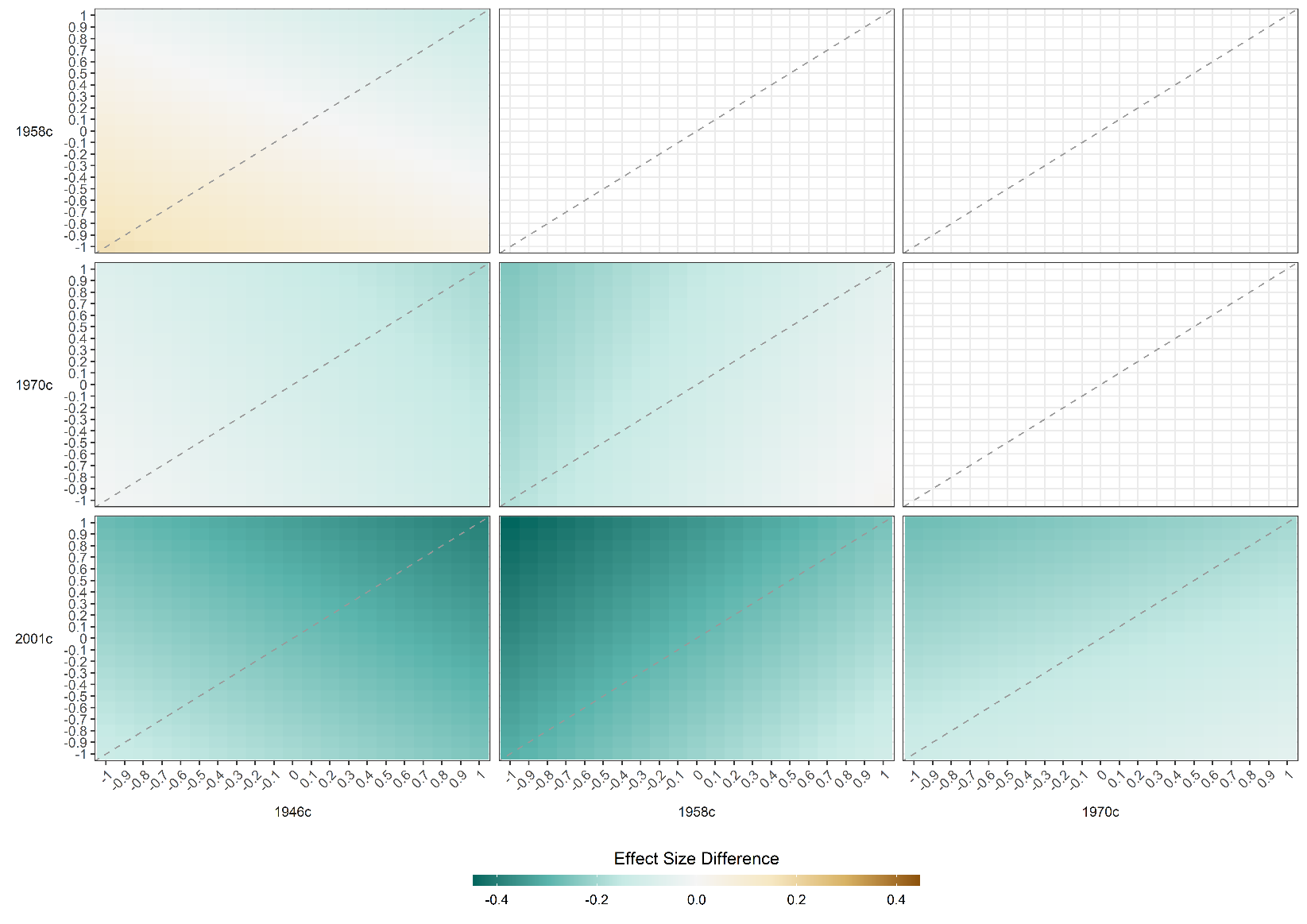

Figure S10: Result of pattern mixture models. Difference in association between height and verbal similarity @ age 11 across cohort. Derived from pooled OLS models (32 imputed datasets) including adjustment for sex. OLS models repeated with imputed height values adjusted by a constant factor given in columns and rows. The colour of each square indicates difference in the association between height and cognition between two cohorts with the coefficient for the cohort on the horizontal subtracted from that on the horizontal (with given adjustment factors used). For instance, in the bottom left panel, the co-ordinate 1, -1 refers to difference in cofficients where +1 SD was added to imputed height values in 1946c and -1 SD was added to imputed height values in 2001c. Height harmonised across cohorts using growth charts from 1990 UK growth study. Cognition scores harmonised using ridit scoring (range 0-1).

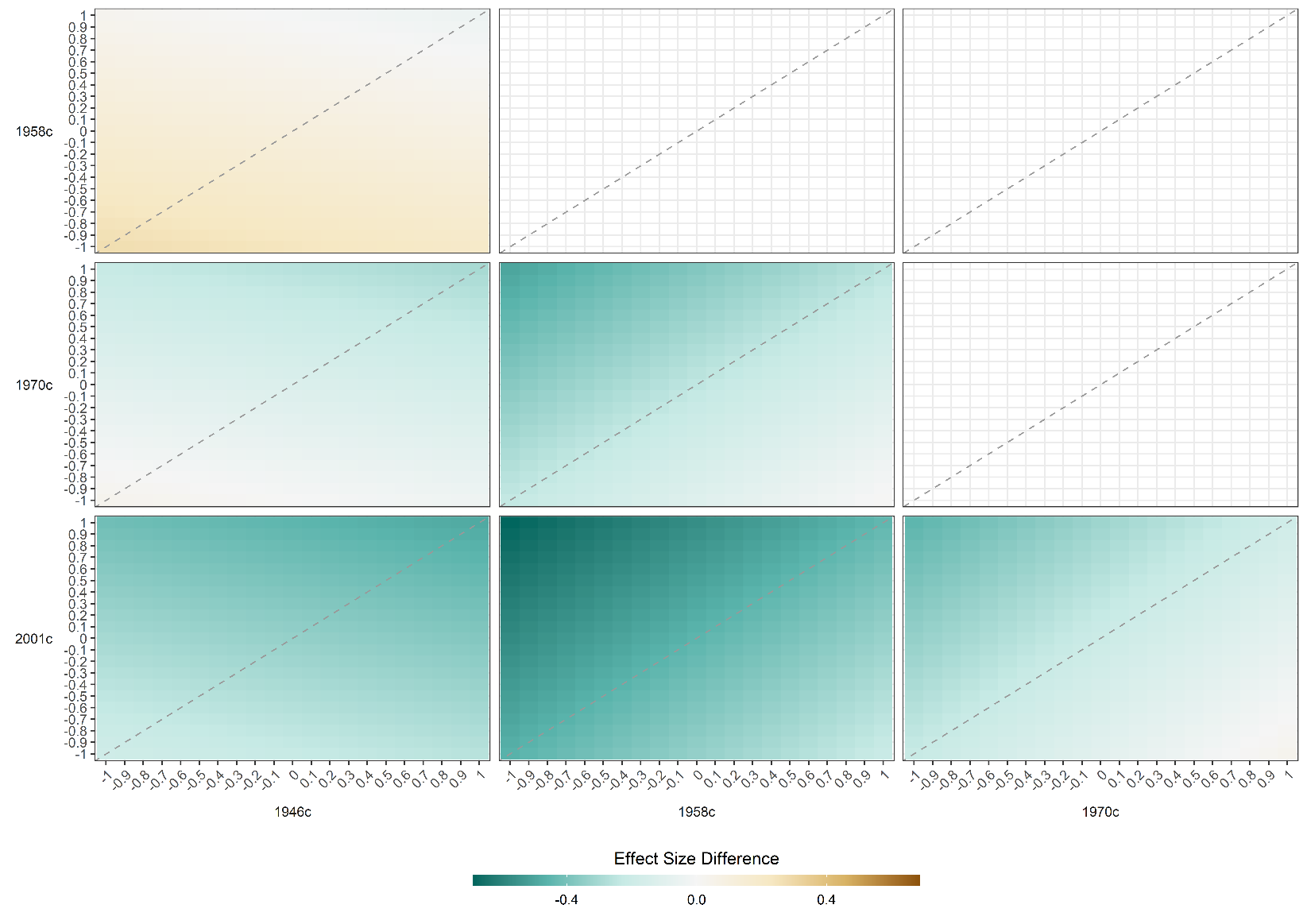

Figure S11: Result of pattern mixture models. Difference in association between height and vocabulary/comprehension @ age 16 across cohort. Derived from pooled OLS models (32 imputed datasets) including adjustment for sex. OLS models repeated with imputed height values adjusted by a constant factor given in columns and rows. The colour of each square indicates difference in the association between height and cognition between two cohorts with the coefficient for the cohort on the horizontal subtracted from that on the horizontal (with given adjustment factors used). For instance, in the bottom left panel, the co-ordinate 1, -1 refers to difference in cofficients where +1 SD was added to imputed height values in 1946c and -1 SD was added to imputed height values in 2001c. Height harmonised across cohorts using growth charts from 1990 UK growth study. Cognition scores harmonised using ridit scoring (range 0-1).

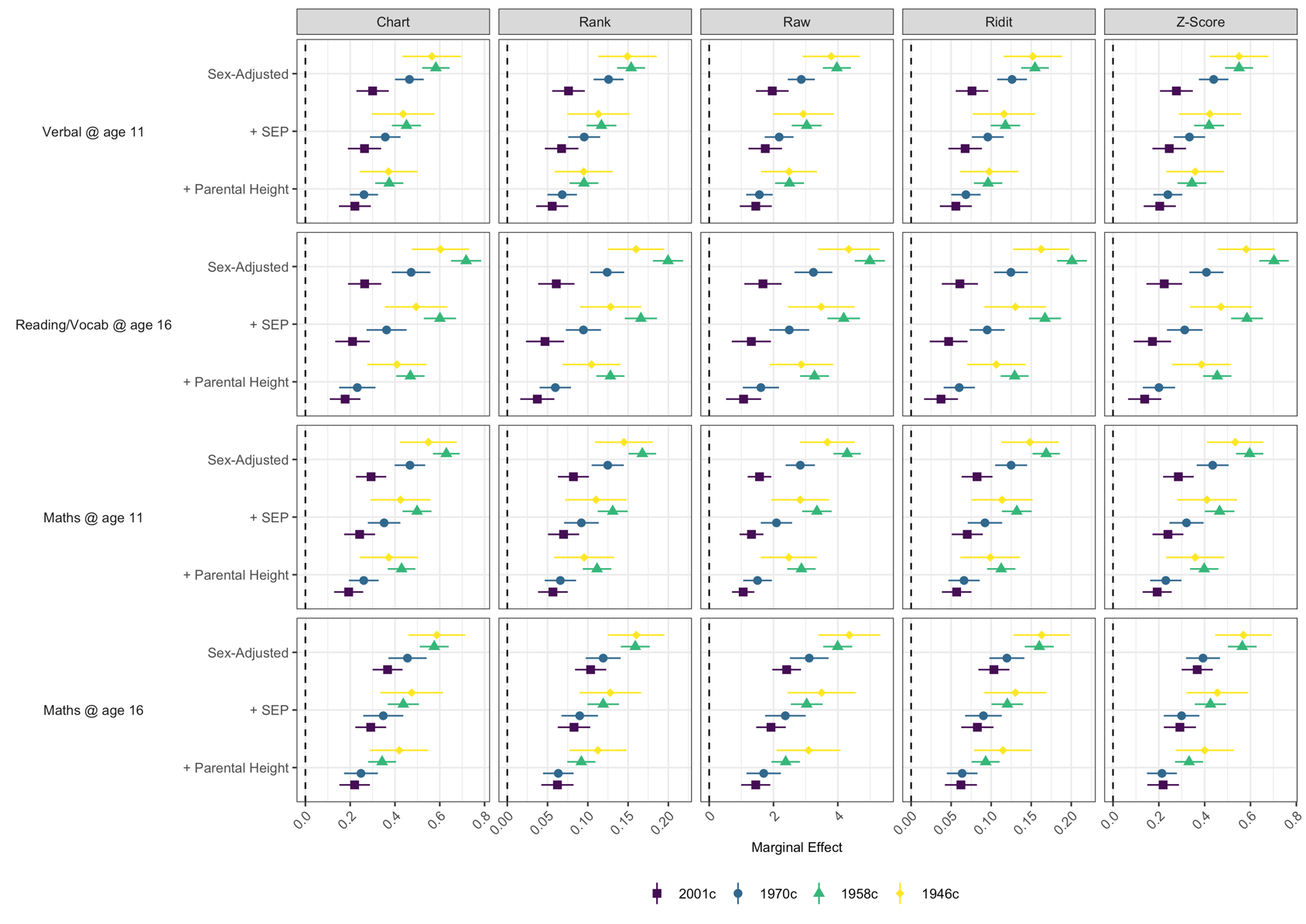

Figure S12: Association between height and cognition by cohort, test (vertical panels)), and procedure used to score height values (horizontal panels). Derived from pooled OLS models (32 imputed datasets), adjusting for (rows) sex; sex, mother’s education and father’s social class (middle panel), and sex, mother’s education, father’s social class, and paternal and maternal height. Height scored using reference centiles from 1990 UK growth study, percentile ranks, raw values, ridit scoring, and Z-scoring in each cohort and sex, respectively. Cognition scores harmonised using ridit scoring (range 0-1).

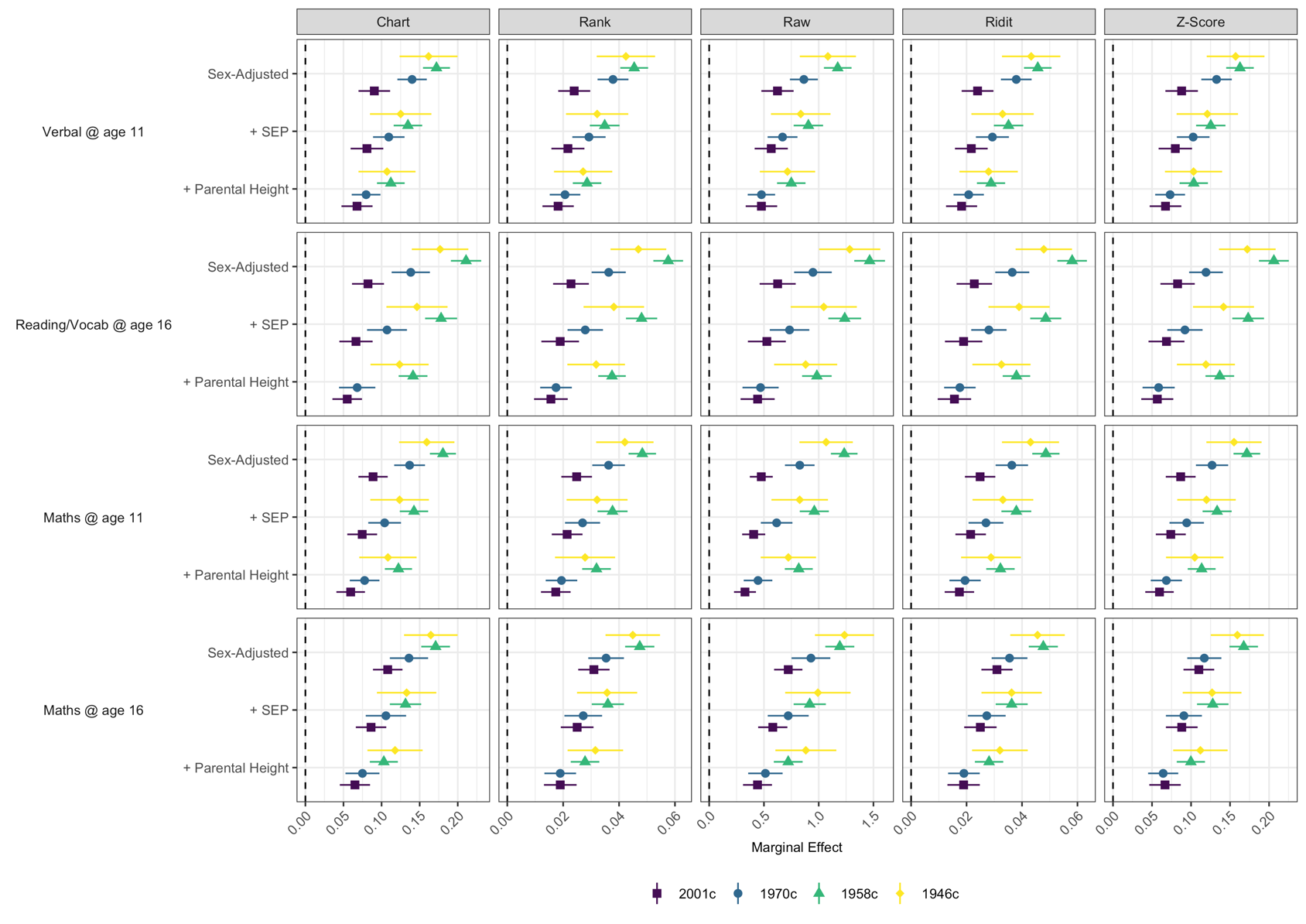

Figure S13: Association between height and cognition by cohort, test (vertical panels)), and procedure used to score height values (horizontal panels). Derived from pooled OLS models (32 imputed datasets), adjusting for (rows) sex; sex, mother’s education and father’s social class (middle panel), and sex, mother’s education, father’s social class, and paternal and maternal height. Height scored using reference centiles from 1990 UK growth study, percentile ranks, raw values, ridit scoring, and Z-scoring in each cohort and sex, respectively. Cognition scores harmonised using Z-scores (mean = 0, SD = 1).

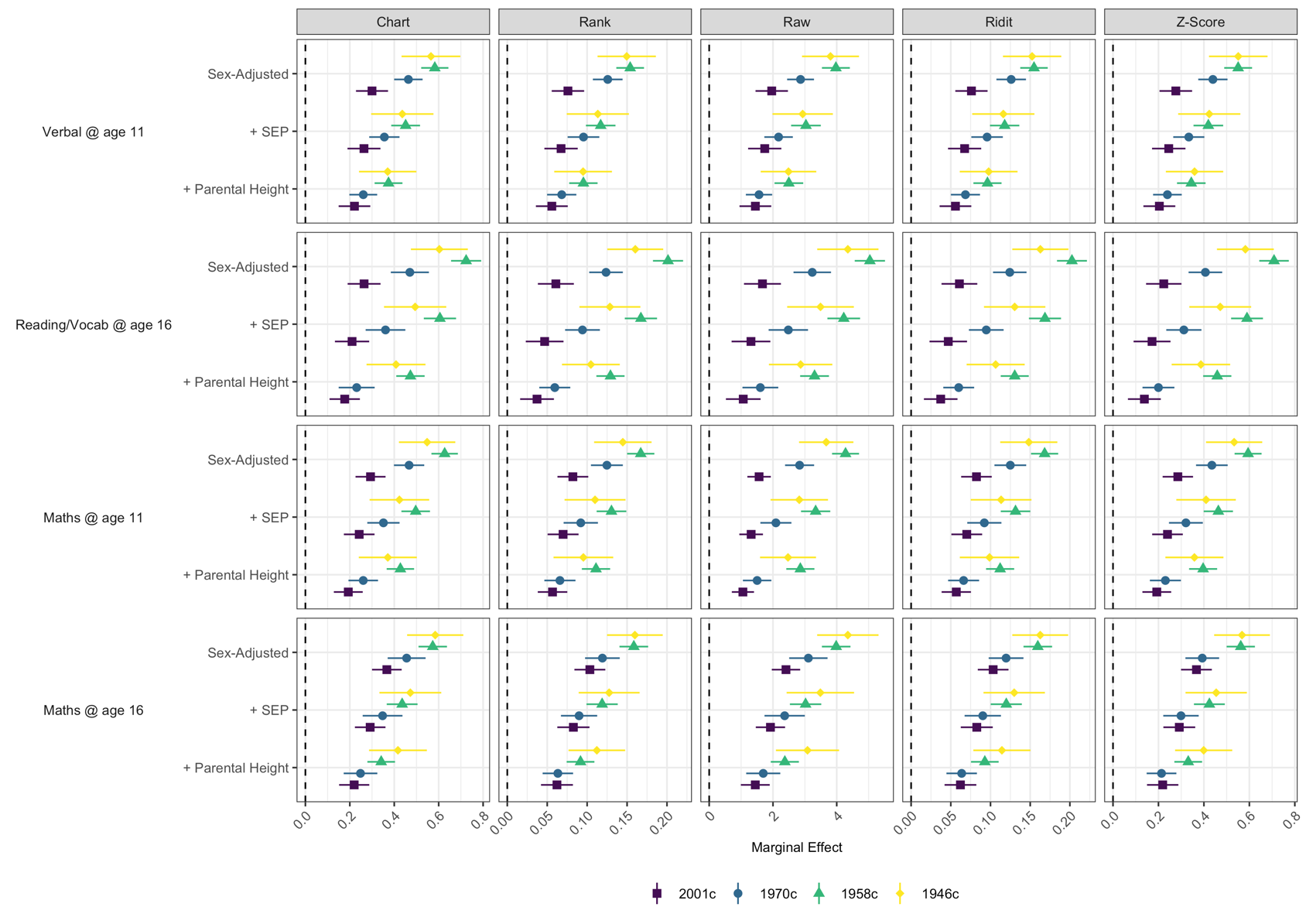

Figure S14: Association between height and cognition by cohort, test (vertical panels)), and procedure used to score height values (horizontal panels). Derived from pooled OLS models (32 imputed datasets), adjusting for (rows) sex; sex, mother’s education and father’s social class (middle panel), and sex, mother’s education, father’s social class, and paternal and maternal height. Height scored using reference centiles from 1990 UK growth study, percentile ranks, raw values, ridit scoring, and Z-scoring in each cohort and sex, respectively. Cognition scores harmonised using percentile ranks (range 0-1).

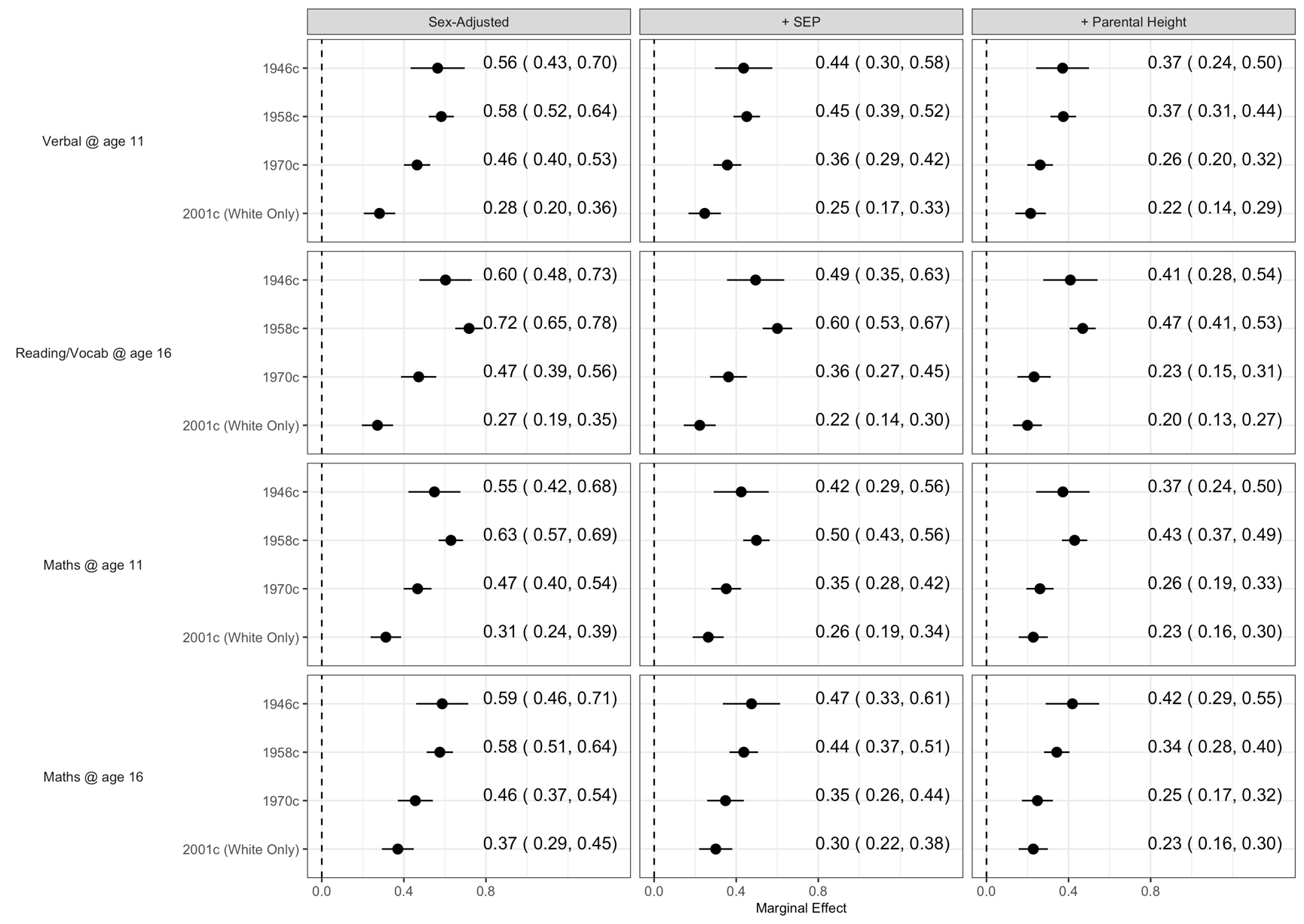

Figure S15: Association between height and cognition by cohort and test. Derived from pooled OLS models (32 imputed datasets), adjusting for sex (left panel); sex, mother’s education and father’s social class (middle panel), and sex, mother’s education, father’s social class, and paternal and maternal height (right panel). Height harmonised across cohorts using growth charts from 1990 UK growth study. Cognition scores harmonised using ridit scoring (range 0-1). 2001c participants limited to individuals of white ethnicity.

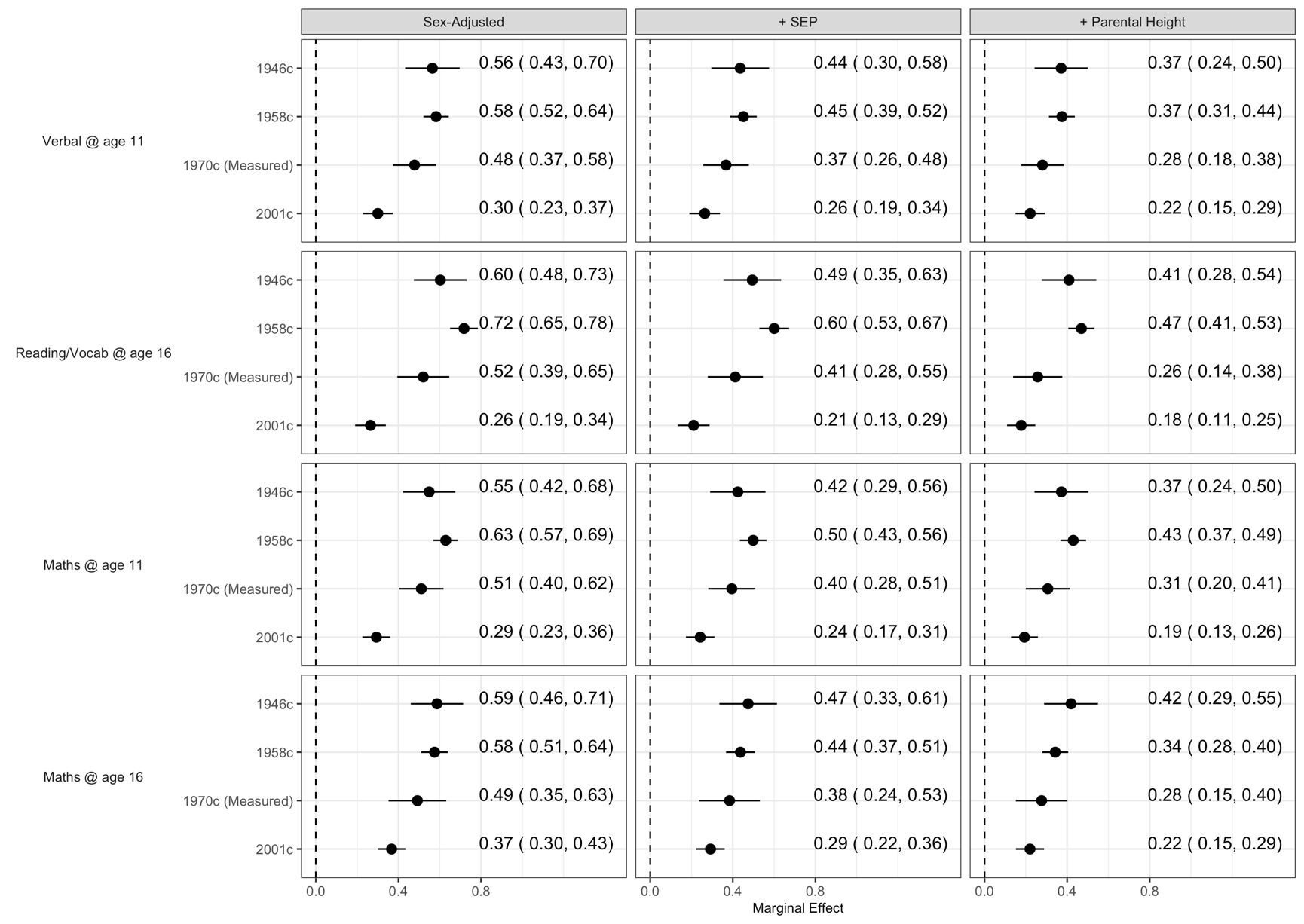

Figure S16: Association between height and cognition by cohort and test. Derived from pooled OLS models (32 imputed datasets), adjusting for sex (left panel); sex, mother’s education and father’s social class (middle panel), and sex, mother’s education, father’s social class, and paternal and maternal height (right panel). Height harmonised across cohorts using growth charts from 1990 UK growth study. Cognition scores harmonised using ridit scoring (range 0-1). 1970c participants limited to individuals with measured height at age 16.
